# Evaluation of antibody serology to determine current helminth and *Plasmodium falciparum* infections in a co-endemic area in Southern Mozambique

**DOI:** 10.1101/2022.01.18.22268971

**Authors:** Rebeca Santano, Rocío Rubio, Berta Grau-Pujol, Valdemiro Escola, Osvaldo Muchisse, Inocência Cuamba, Marta Vidal, Gemma Ruiz-Olalla, Ruth Aguilar, Javier Gandasegui, Maria Demontis, Jose Carlos Jamine, Anélsio Cossa, Charfudin Sacoor, Jorge Cano, Luis Izquierdo, Chetan E. Chitnis, Ross L Coppel, Virander Chauhan, David Cavanagh, Sheetij Dutta, Evelina Angov, Lisette van Lieshout, Bin Zhan, José Muñoz, Carlota Dobaño, Gemma Moncunill

**Affiliations:** ISGlobal, Hospital Clínic - Universitat de Barcelona, Barcelona, Spain; Centro de Investigação em Saúde de Manhiça (CISM), Maputo, Mozambique; Fundación Mundo Sano, Argentina; Department of Parasitology, Centre of Infectious Diseases, Leiden University Medical Centre (LUMC), Leiden, The Netherlands; Communicable and Non-communicable Diseases Cluster (UCN), WHO Regional Office for Africa, Brazzaville, Republic of Congo; CIBER de Enfermedades Infecciosas, Madrid, Spain; Malaria Parasite Biology and Vaccines, Department of Parasites & Insect Vectors, Institut Pasteur, Paris, France; Department of Microbiology, Faculty of Medicine, Nursing and Health Sciences Monash University, Melbourne, Australia; Malaria Group, International Centre for Genetic Engineering and Biotechnology (ICGEB), New Delhi, India; Institute of Immunology and Infection Research, University of Edinburgh, Edinburgh, UK; Walter Reed Army Institute of Research (WRAIR), Maryland, USA; Baylor College of Medicine (BCM), Texas, USA

## Abstract

**Background:** Soil-transmitted helminths (STH), *Schistosoma* spp. and *Plasmodium falciparum* are parasites of major public health importance and co-endemic in many sub-Saharan African countries. Management of these infections requires detection and treatment of infected people and evaluation of large-scale measures implemented. Diagnostic tools are available but their low sensitivity, especially for low intensity helminth infections, leaves room for improvement. Antibody serology could be a useful approach thanks to its potential to detect both current infection and past exposure.

**Methodology:** We evaluated total IgE responses and specific-IgG levels to 9 antigens from STH, 2 from *Schistosoma* spp., and 16 from *P. falciparum*, as potential markers of current infection in a population of children and adults from Southern Mozambique (N = 715). Antibody responses were measured by quantitative suspension array Luminex technology and their performance was evaluated by ROC curve analysis using microscopic and molecular detection of infections as reference.

**Principal findings:** IgG against the combination of EXP1, AMA1 and MSP2 (*P. falciparum*) in children and NIE (S*trongyloides stercoralis*) in adults and children had the highest accuracies (AUC = 0.942 and AUC = 0.872, respectively) as markers of current infection. IgG against the combination of MEA and Sm25 (*Schistosoma* spp.) were also reliable markers of current infection (AUC = 0.779). In addition, IgG seropositivity against 20 out of the 27 antigens in the panel differentiated the seropositive endemic population from the non-endemic population, suggesting a possible role as markers of exposure.

**Conclusions:** We provided evidence for the utility of antibody serology to detect current infection with parasites causing tropical diseases in endemic populations. In addition, most of the markers could be used as markers of exposure. We also showed the feasibility of measuring antibody serology with a platform that allows the integration of control and elimination programs for different pathogens.

**AUTHOR SUMMARY:** Parasitic worms and *Plasmodium falciparum*, the causal agent of malaria, are among the most relevant parasitic diseases of our time and efforts are under way for their control and, ultimately, elimination. An accurate diagnosis is relevant for case management, but also allows calculating the prevalence and evaluating the effectiveness of treatment and control measures. Unfortunately, current diagnostic methods for parasitic worms are not optimal and many infections remain undetected. As for *P. falciparum*, current diagnostic techniques are satisfactory but do not allow for ascertaining exposure, which is relevant for evaluating control measures. Here we investigated the utility of measuring antibodies to these parasites as a diagnostic method. Our results indicate that it is possible to detect current infection with parasitic worms and *P. falciparum* using antibody detection with a moderate to high accuracy. We also show that antibodies could distinguish a population from Southern Mozambique, where these infections are prevalent, from a Spanish population never exposed to those parasites. Importantly, we used a platform that allows for the simultaneous detection of immunoglobulins to different parasites, which would be extremely useful as a tool to integrate control and elimination programs for several pathogens.

## INTRODUCTION

Soil-transmitted helminths (STH) and *Schistosoma* spp. are amongst the most relevant neglected tropical diseases (NTDs), affecting 1.5 billion and 236.6 million people worldwide, respectively (1, 2). STH infections are caused by the parasitic worms *Ascaris lumbricoides*, *Ancylostoma duodenale*, *Necator americanus*, *Trichuris trichiura* and *Strongyloides stercoralis*. Malaria, caused by parasites of the genus *Plasmodium*, results in a high morbidity and mortality, with 241 million cases and 627,000 deaths in 2020 (3). Both types of parasitic diseases overlap geographically, especially in areas of sub-Saharan Africa (4).

Microscopic observation of eggs or larvae is the recommended procedure for the diagnosis of STH and *Schistosoma* spp. infections. However, it requires well-trained personnel, is laborious and time consuming. It is also dependent upon the intermittent shedding of eggs or larvae, which requires collection of samples from different days for a correct detection (5–8). On top of that, the impact of the low sensitivity of microscopy is exacerbated with lower intensity and prevalence of infections, which are likely after decades of Mass Drug Administration (MDA) to control these infections (9).

The diagnosis of *Plasmodium* spp. is most commonly done by the detection of parasites, their genetic material or antigens in blood by microscopy, quantitative real-time polymerase chain reaction (qPCR) or rapid diagnostic tests (RDT), respectively (10). These methods also face challenges, such as low parasitemia levels that remain undetected in the case of microscopy and RDT, the need for well-trained personnel for microscopy or the required equipment in low-income settings for qPCR.

Antibody serology could be a useful tool for detection of current infection but also exposure, which is not captured by any of the diagnostic methods above. For helminths, antibody serology has a particularly important role in the diagnosis of *S. stercoralis* infection, since conventional methods have a very low sensitivity and specific antibodies generally decrease 6 months after treatment, which helps detect current and recent infections (11–13). In fact, antibody serology against the NIE antigen from *S. stercoralis* has been used to monitor MDA programs thanks to the rapid decay of antibodies after clearance of infection (14, 15). For other species, antibody serology is less common and not routinely implemented. However, there is evidence of their potential. For example, a study has shown that increased levels of immunoglobulin G4 (IgG4) against the *Ascaris* spp. antigen AsHb reflected recent exposure to the parasite (16), and another study reported moderate accuracy of IgG against *N. americanus* proteins in detecting current infection (17). In the case of *Schistosoma* spp., there are commercial kits that might be useful for the diagnosis of imported schistosomiasis in individuals from non-endemic or low-endemic areas, while in high-to moderate-endemic areas they have proven limited utility (18, 19). For malaria, serology has mostly been used to measure exposure and determine malaria endemicity, transmission intensity and to monitor interventions (20, 21). In fact, this ability of antibodies to capture exposure even in the absence of current infection has been exploited as an useful tool in control and elimination programs for other infections to monitor changes in transmission, such as the elimination programs of lymphatic filariasis (22), onchocerciasis (23) and trachoma (24).

Identification of patent helminth infection and *Plasmodium* spp. parasitemia by serology can be challenging, particularly in endemic areas, due to the generally long-lasting antibody responses and repeated exposure (21, 25). However, it might be useful in patients who have been in endemic areas for the first time, for less prevalent infections, or in cases of very low parasite burden (26, 27), for which highly sensitive diagnostic tests are required.

Increasing evidence indicates that the potent immunomodulatory effects of helminths might have effects on the immune response to *P. falciparum* (4). Moreover, we have recently shown that this interaction might be bidirectional, since it was observed that exposure to or co-infection with helminths and *P. falciparum* was associated with an increase in antibody responses to antigens from both pathogens (28). Given the reported bidirectional impact on the immune responses to both parasitic infections, it is relevant to study antibody serological diagnostic methods in a context where co-infections might occur and affect the accuracy of the diagnosis. In this study we have evaluated total IgE and antigen-specific IgGs against a wide panel of antigens from helminths (STH, *Schistosoma* spp.) and *P. falciparum* parasitic infections as potential markers of current infection.

## METHODS

### Study samples

Stool, urine, and blood samples were collected from children (5-15 years old) (N= 363) and adults (>15 years old) (N= 352) from a malaria (29) and helminth (30) endemic area in Southern Mozambique (N = 715) in the context of a community-based, case-cohort study (MARS), described in detail elsewhere (30). Serum samples from Spanish adult donors were used as unexposed negative controls (NC) (N = 50) in the serological assays. They represent individuals from a non-endemic area who had never travelled to any countries endemic for the infections addressed in the study.

### Ethics

The study was carried out according to the principles of the Declaration of Helsinki. Written informed consent from individuals older than 18 years old who were willing to participate was obtained. In the case individuals younger than 18 years old, their parents or guardians provided a written informed consent. Additionally, participants from aged 15 to 17 also gave informed assent. For illiterate individuals, informed consent was conducted in presence of a literate witness independent from study. Samples analyzed in this study received ethical clearance for immunological evaluation and/or inclusion as controls in immunoassays. The protocols and informed consent forms were approved by the Institutional Review Board at Hospital Clínic in Barcelona (Ref.: CEIC-7455) and the National Bioethics Committee for Health in Mozambique (Ref.: 517/CNBS/17).

### Detection of helminth and *P. falciparum* infections by the reference methods

Helminth infections were detected by microscopic and molecular diagnosis at the Manhiça Health Research Centre (CISM) (Mozambique) and Leiden University Medical Center (The Netherlands), respectively.

Microscopic diagnosis of *N. americanus*, *A. duodenale*, *T. trichiura, A. lumbricoides* and *S. mansoni* was performed by search of eggs in two stool samples from two consecutive days using duplicate Kato-Katz thick smear technique and Telemann technique (31, 32). *S. stercoralis* was detected in two stool samples from two consecutive days using the Telemann technique (32). *S. haematobium* infection was detected by microscopy after urine filtration (31). Individuals were considered to be positive for helminth infection by copromicroscopy if they had a positive result in at least one of the stool samples (30).

Molecular diagnosis was also performed in one stool sample per participant by multiplex semi-qPCR, as described in detail elsewhere (30). Two different multiplex detection panels were used; panel 1 targeted *Schistosoma* spp. and *T. trichiura*, and panel 2 targeted *A. duodenale*, *N. americanus, A. lumbricoides* and *S. stercoralis*. Of note, qPCR to detect helminth infections was not performed in 25 out of the 715 samples from the endemic population.

*P. falciparum* infection was determined at the Barcelona Institute for Global Health (ISGlobal) (Spain) by 18S ribosomal RNA gene detection through qPCR from dried blood spot (DBS) samples as previously described (28).

### Serological assays

A detailed explanation of the antigen panel and serum elution procedure from DBS used for this analysis is described in Santano *et al*. 2021 (28). Eluted serum was used to measure total IgE and antigen-specific IgG by quantitative suspension array technology (qSAT) from Luminex against a panel of 27 antigens: 16 from *P. falciparum* (α-gal, CelTOS, SSP2, LSA1, EXP1, AMA1, EBA175, MSP1 block 2, MSP1_42_, MSP2, MSP3, MSP5, P41, RH1, RH5 and PTRAMP and 11 from helminths (hookworm [NaGST1, NaAPR1, NaSAA2, AyCp2], *Trichuris* spp. [TmWAP, Tm16], *Ascaris* spp. [As16, As37], *S. stercoralis* [NIE] and *Schistosoma* spp. [MEA, Sm25]). The antigens and a capture anti-human IgE (Mouse anti-Human IgE Fc; ref. ab99834, Abcam PLC, Cambridge, UK) were coupled to magnetic MagPlex® 6.5μm COOH-microspheres (Luminex Corporation, Austin, USA). The antigen-coupled beads were used in multiplex while the anti-human IgE-coupled beads were used in singleplex. Coupled beads were incubated with test and control serum samples, in black 96-well μClear® flat bottom plates at 4 °C overnight (ON) in a shaker at 600 rpm. For the antigen-specific IgG quantification we assayed the samples at 1:100 and 1:2000 dilutions, and for total IgE quantification we assayed the samples at 1:50 dilution. After the ON incubation beads were washed three times with PBS-0.05% Tween20, using a manual magnetic washer platform (ref. 40-285, Bio-Rad, Hercules, USA), and the biotinylated secondary anti-human IgG (ref. B1140-1ML, MilliporeSigma, St. Louis, USA)] and anti-human IgE (ref. A18803, Invitrogen, Waltham, USA) antibodies were added at 1:1250 and 1:100 dilutions, respectively, and incubated for 45 min at room temperature (RT) shaking at 600 rpm. Then, beads were washed three times and streptavidin-R-phycoerythrin (ref. 42250-1ML, MilliporeSigma, St. Louis, USA) was added at 1:1000 dilution and incubated at RT for 30 min at 600 rpm. Finally, beads were washed, resuspended, and at least 50 beads per analyte and sample were acquired in a FlexMap 3D xMAP® instrument (Luminex Corporation, Austin, USA). Crude median fluorescent intensities (MFI) and background fluorescence from blank wells were exported using the xPONENT software v.4.3 (Luminex Corporation, Austin, USA).

### Data analysis

To determine the seropositivity and perform the statistical analysis, normalized and dilution corrected antibody data were log_10_-transformed. Detailed information on the processing of data can be found in Santano *et al*. 2021 (28).

The receiver operating characteristic (ROC) curves and their corresponding areas under the curve (AUC) for each antibody response were built with the R CRAN package pROC (33) using the log_10_-transformed MFIs as predictor and the microscopic, molecular or combination of both diagnosis as response variable. Since antibodies are acquired over time (28), we also performed the analysis stratified by age group (children and adults). In addition, to eliminate the possible interference of helminths and *P. falciparum* co-infections with the performance of the assessed markers, we estimated additional ROC curves and AUC analyses where the log_10_ MFIs from samples with only one infection (cases) or no infection (controls) diagnosed by the corresponding method were used as predictors, and the microscopic, molecular or combination of both diagnosis as response variables. Finally, we also evaluated the performance of combined antibody serological markers for those parasites for which more than one antigen was available. To do so, we built logistic regression models using the R function “glm” with the combination of log_10_-transformed MFIs as predictor variable and the microscopic, molecular or combination of both diagnosis as response variable. The predicted values fitted by those models were then used to build the ROC curves and calculate the corresponding AUCs. For those parasites with only two antigens in the panel (*Ascaris* spp., *Trichuris* spp. and *Schistosoma* spp.), we performed the analysis with the combination of those two antigen-specific IgG levels. For those with more than two antigens (*P. falciparum* and hookworm) we performed all the possible combinations and selected the one that gave the best AUC with the lowest number of predictors. In the case of *P. falciparum* antigens, we included only the top performers to calculate the combinations, whether for the whole population or stratified by age. For the analysis of antigen-specific IgG, MFI data from the 715 samples from endemic individuals and the 50 samples from Spanish donors were used. In the case of total IgE only MFI data from endemic individuals were used, since there are other IgE-inducing pathologies, such us allergy, which are prevalent in Spain and render these samples not useful as negative controls.

Five seropositivity cutoffs were compared to evidence the options that can be used in scenarios requiring prioritization of specificity, sensitivity or a more balanced performance, and to contrast the performance of cutoffs calculated with data from non-endemic or endemic populations. The Expectation-Maximization (EM) cutoff was estimated using Gaussian Mixture Models for each antigen-specific IgG and total IgE in the study population. Two models, estimated by the EM algorithm, were fitted assuming either equal or unequal variances, and each of those two models were built with two clusters for classification, which were meant to represent the distribution of seropositive and seronegative samples in the endemic population. For each antigen, classification of samples was done using the model showing the optimal Bayesian Information Criterion (BIC). The models were fitted using the R CRAN package mclust (34). The NC cutoff was calculated as the mean plus 3 standard deviations of the log_10_-transformed MFIs from the Spanish donors for each antigen-specific IgG. Finally, the ROC curves were used to generate three different cutoffs. The first one prioritized a minimum specificity of 95% (95SP) and the maximum sensitivity within the possible cutoffs. The second one prioritized a minimum sensitivity of 95% (95SE) and the maximum specificity within the possible cutoffs. The third one prioritized the maximum Youden’s index (YI), which is the cutoff with the maximum distance from the line of identity.

We then evaluated whether having an infection with any of the parasites detected in the study had an effect on the specific IgG levels and therefore influenced the false positive or false negative rates for a given seropositivity cutoff. To do so, for each antigen in the panel we selected the individuals who had a negative diagnosis for the corresponding species (i.e.: for NIE [from *S. stercoralis*] we selected all individuals with a negative *S. stercoralis* diagnosis). Among these individuals, we selected those who had only one single infection (to avoid interferences from co-infections) or no infection diagnosed. Finally, we compared the antibody levels for each species to the levels of those with no infection diagnosed. Pairwise comparisons were done by Wilcoxon rank-sum test and Holm adjustment for multiple testing. P values < 0.05 were considered statistically significant.

All data processing and statistical analyses were performed using the statistical software R version 4.0.3. R CRAN packages used to manage data, generate tables and plots were tidyverse (35), ggpubr (36) and ggbeeswarm (37).

## RESULTS

### Antibody serological markers of current helminth infection

A good serological marker of current infection should be able to distinguish those currently infected from those not infected although previously exposed. The ROC curves analyses and their corresponding AUC showed that IgG levels against most of the helminth antigens in the panel had a low (0.50 ≤ AUC > 0.70) or null accuracy (0.50 < AUC) (38) in detecting current helminth infection as defined by microscopy, qPCR or their combination (**Fig 1A** and **Table S1**). However, IgG levels against some antigens showed a moderate accuracy (0.70 ≤ AUC > 0.90) in detecting current infection. Namely, NIE from *S. stercoralis* had an AUC = 0.872 using qPCR as a reference, while the AUCs using microscopy or both techniques were slightly lower (**Fig 1A** and **Fig 2**). However, the number of *S. stercoralis* infections detected by microscopy was very low (N = 8), probably giving unreliable results. MEA and Sm25 from *Schistosoma* spp. had AUC = 0.746 and AUC = 0.741, respectively, using *S. mansoni* microscopy data as reference (**Fig 1A** and **Fig 2**). Sm25 also showed moderate AUC when using *Schistosoma* spp. microscopy (AUC = 0.721), *Schistosoma* spp. qPCR (AUC = 0.712) or *S. haematobium* microscopy (AUC = 0.711) as references (**Fig 1A** and **Fig 2**). The remaining cases had all AUC < 0.700 (**Fig 1A** and **Table S1**). When all specific-IgG responses were combined for helminths with more than one antigen in the panel, we observed a slight improvement in the AUCs for *Schistosoma* spp. (**Fig S1A**) but no improvement for the rest (data not shown).

**Fig 1.**
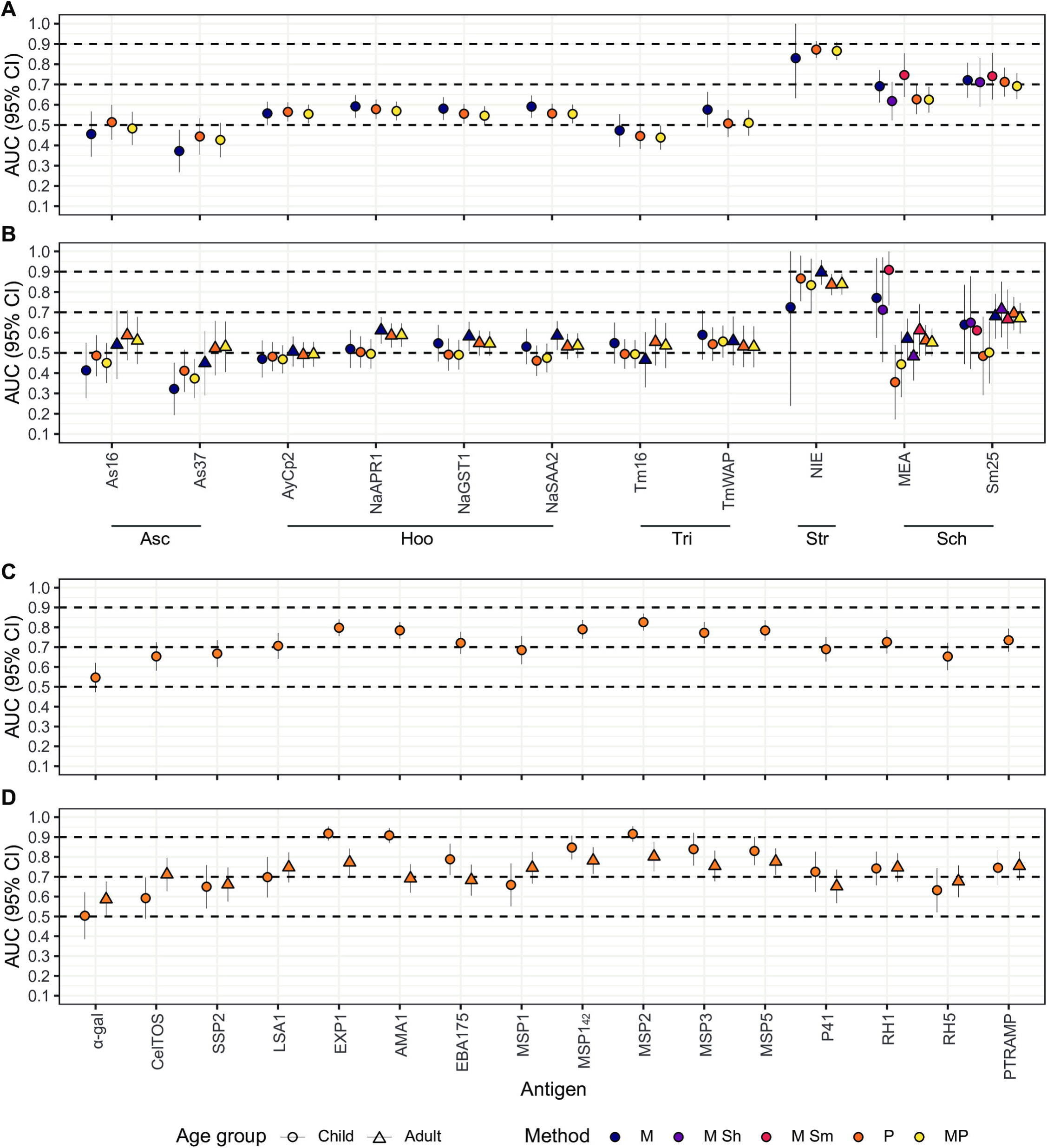
Areas under the curve from receiver operating characteristic curve analysis. **A** and **B** show the performance of helminth-specific IgG levels for the whole population or stratified by age, respectively. **C** and **D** show the performance of malaria-specific IgG levels for the whole population or stratified by age, respectively. Dashed lines represent the accuracy thresholds: null if AUC < 0.5, low if 0.5 ≤ AUC > 0.7, moderate if 0.7 ≤ AUC > 0.9 and high if AUC ≥ 0.9. AUC: Area Under the Curve; CI: Confidence Interval; M: Microscopy; P: qPCR; MP: Microscopy and qPCR combined. Asc: *Ascaris* spp.; Hoo: Hookworm; Tri: *Trichuris* spp.; Str: *Strongyloides stercoralis*; Sch: *Schistosoma* spp. Complementary information for this figure can be found in supplementary Table S1 and S2.

**Fig 2.**
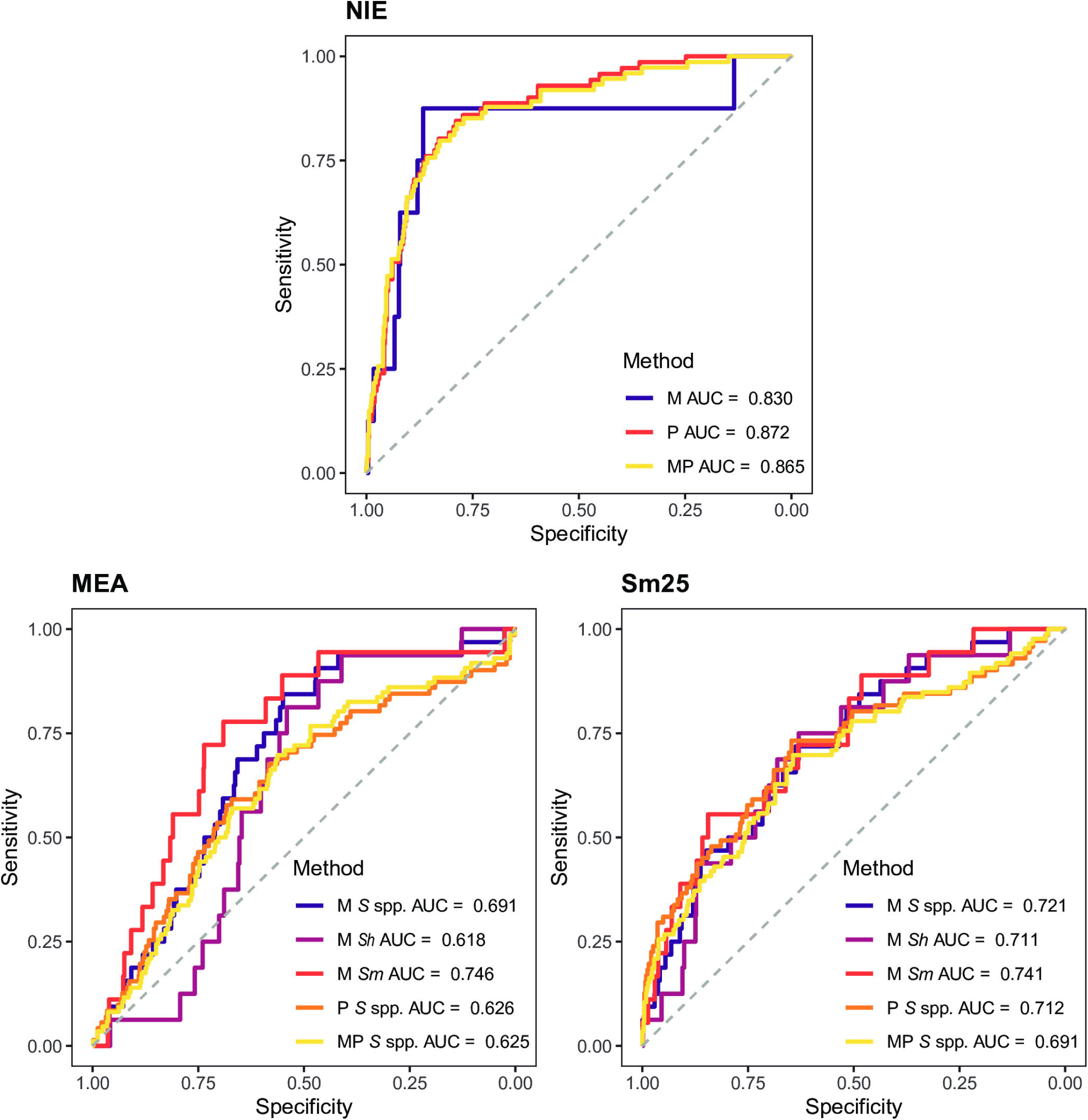
Receiver operating characteristic curves and their corresponding areas under the curve from the best performing IgG responses in detecting current helminths infection. IgG responses (log_10_-transformed median fluorescence intensity [MFI] levels) against *S. stercoralis* (NIE) and *Schistosoma* spp. (MEA and Sm25) antigens in serum samples from 715 endemic individuals and 50 Spanish donors were used as predictor variables. Microscopic (M), qPCR (P) or the combination of both (MP) diagnoses were used as response variables. *S* spp.: *Schistosoma* spp.; *Sh*: *Schistosoma haematobium*; *Sm*: *Schistosoma mansoni*. AUC: Area Under the Curve.

After stratifying by the age group, the performance for NIE improved for adults when using microscopy as reference, but the cases were very few (N = 5) to generate reliable results (**Fig 1B** and **Table S1**). On the other hand, the results using qPCR or qPCR combined with microscopy as references were only slightly better in children compared to adults or equal, respectively, but lower than before the age stratification (**Fig 1B** and **Table S1**). As for *Schistosoma* spp., moderate and high AUC values were higher than in the non-stratified analysis, but the children (N < 5) and adult (N = 11) sample size after stratification was much reduced in those cases (**Fig 1B** and **Table S1**).

Finally, the analysis removing all individuals with other infections besides the one being evaluated (for cases) and any infection at all (for controls), revealed that the AUC reached excellent levels for NIE (AUC > 0.9), although the sample size of cases by microscopy was very small (N = 3) (**Table S3**).

Total IgE levels were also evaluated as a proxy of current helminth infection. They showed low accuracy for most infections, but moderate accuracy in detecting infections by *Schistosoma* spp. (AUC = 0.715), *S. mansoni* (AUC = 0.728), *S. haematobium* (AUC = 0.712) and hookworm (AUC = 0.700) as defined by microscopy (**Fig 3A**, **Fig 4** and **Table S1**). When infections were detected by qPCR or qPCR combined with microscopy, AUC were slightly lower. Total IgE also had a moderate accuracy for *S. stercoralis* (AUC = 0.722), but the sample size of cases diagnosed by microscopy was very small (N=8, **Fig 3A** and **Table S1**). After stratifying by age group, the AUC for hookworm and *Schistosoma* spp. infections either did not improve or were only slightly better in children than adults (**Fig 3B** and **Table S1**). Finally, the analysis removing all individuals with other infections besides the one being evaluated (for cases) or any infection at all (for controls), showed that total IgE had an AUC = 0.707 using microscopic diagnosis of hookworm as reference (**Table S3**). Total IgE using microscopic diagnosis of *S. mansoni* as reference had an AUC = 0.741, but the sample size of controls was small (**Table S3**).

**Fig 3.**
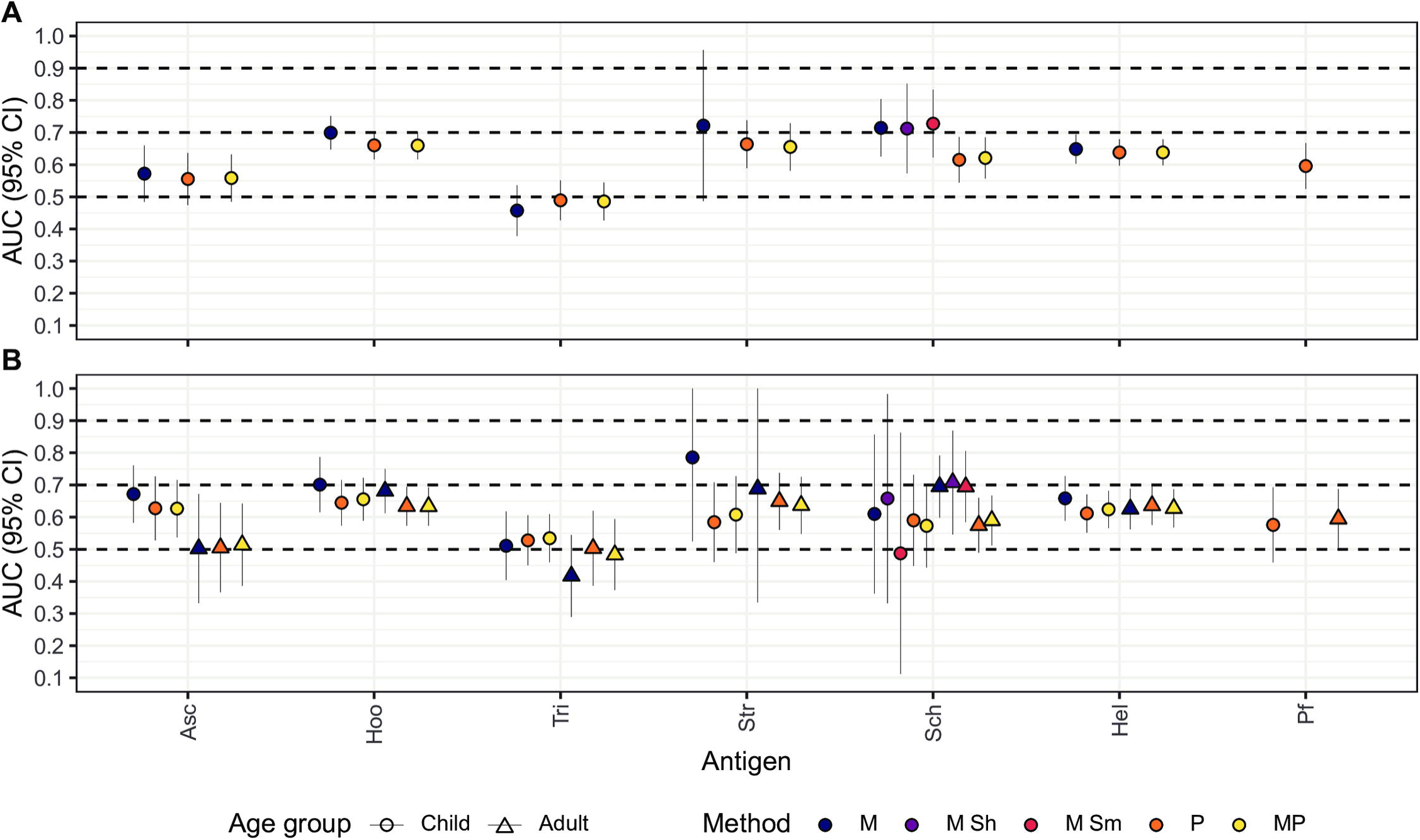
Areas under the curve from receiver operating characteristic curve analysis for total IgE. The performance of total IgE levels to detect helminth or *Plasmodium falciparum* infections in the whole population is shown in **A** and stratified by age in **B**. Dashed lines represent the accuracy thresholds: null if AUC < 0.5, low if 0.5 ≤ AUC > 0.7, moderate if 0.7 ≤ AUC > 0.9 and high if AUC ≥ 0.9. AUC: Area Under the Curve; CI: Confidence Interval; M: Microscopy; P: qPCR; MP: Microscopy and qPCR combined. Asc: *Ascaris* spp.; Hoo: Hookworm; Tri: *Trichuris* spp.; Str: *Strongyloides stercoralis*; Sch: *Schistosoma* spp. Hel: Helminths; Pf: *Plasmodium falciparum.* Complementary information for this figure can be found in supplementary Table S1 and S2.

**Fig 4.**
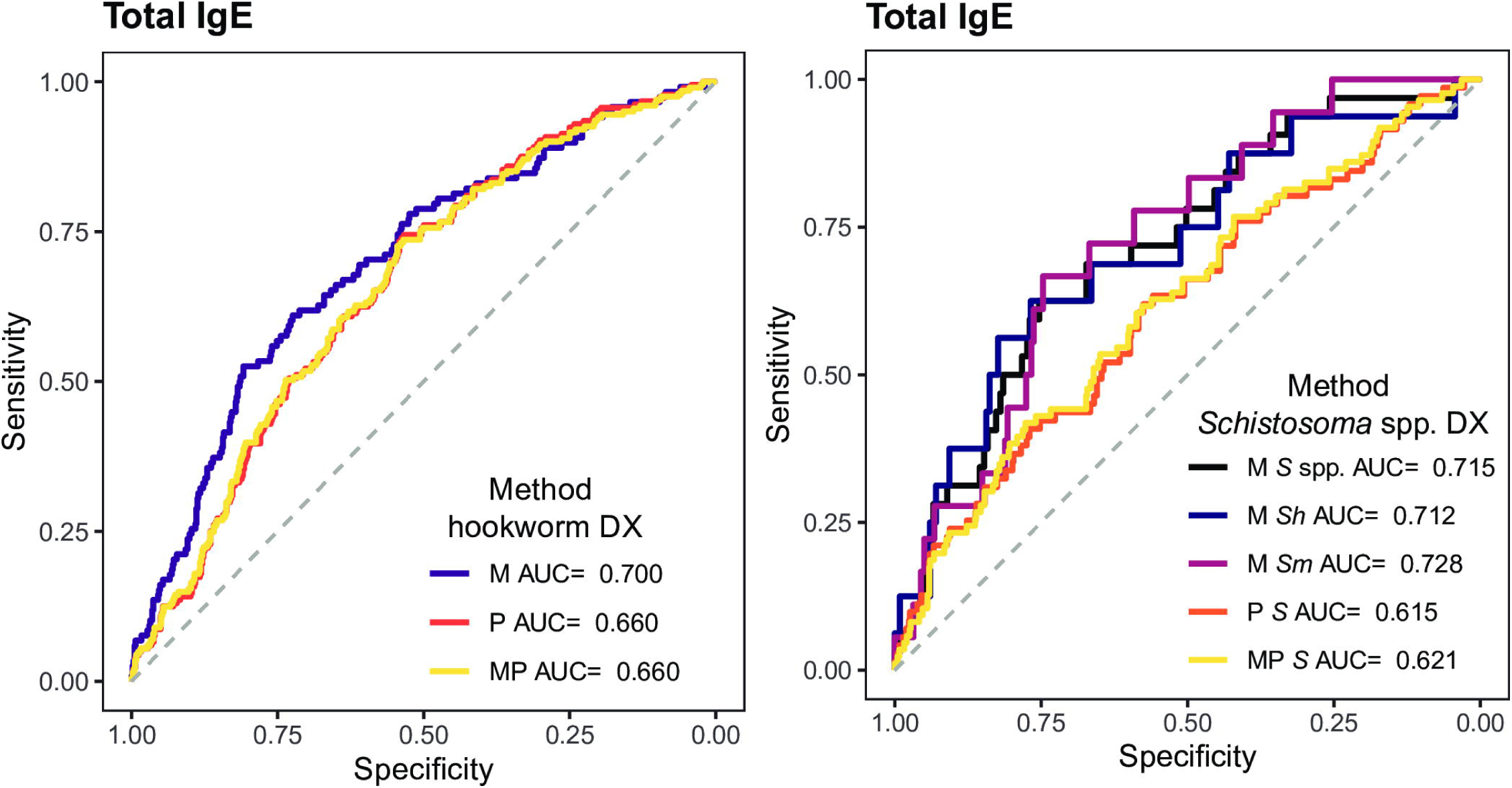
Receiver operating characteristic curves and their corresponding areas under the curve from the best performing total IgE responses in detecting current hookworm and *Schistosoma* spp. infection. Total IgE responses (log_10_-transformed median fluorescence intensity levels) in serum samples from 715 endemic individuals were used as predictor variable. Microscopic (M), qPCR (P) or the combination of both (MP) diagnoses for hookworm and *Schistosoma* spp. were used as response variables. DX: diagnosis; *S* spp.: *Schistosoma* spp.; *Sh*: *Schistosoma haematobium*; *Sm*: *Schistosoma mansoni*; AUC: Area Under the Curve.

### Antibody serological markers of current *P. falciparum* infection

IgG levels against most *P. falciparum* antigens, including MSP2 (AUC = 0.826), EXP1 (AUC = 0.798), MSP1_42_ (AUC = 0.790), AMA1 (AUC = 0.785), MSP5 (AUC = 0.784), MSP3 (AUC = 0.772), PTRAMP (AUC = 0.735), RH1 (AUC = 0.726), EBA175 (AUC = 0.721) and LSA1 (AUC = 0.707) showed a moderate accuracy in detecting *P. falciparum* infection as defined by qPCR diagnosis in asymptomatic individuals (**Fig 1C**, **Fig 5A** and **Table S2**). IgG levels against the remaining *P. falciparum* antigens of the panel had low accuracy in detecting current *P. falciparum* infection (**Fig 1C** and **Table S2**). The top performers mentioned above were used to calculate all possible combinations and build the regression models. The combination that yielded the highest AUC with the simplest formula was AMA1 + EBA175 + MSP1_42_ + MSP2 + MSP3 + MSP5 with an AUC = 0.853 (**Fig S1B**), only slightly better than the best performer MSP2.

**Fig 5.**
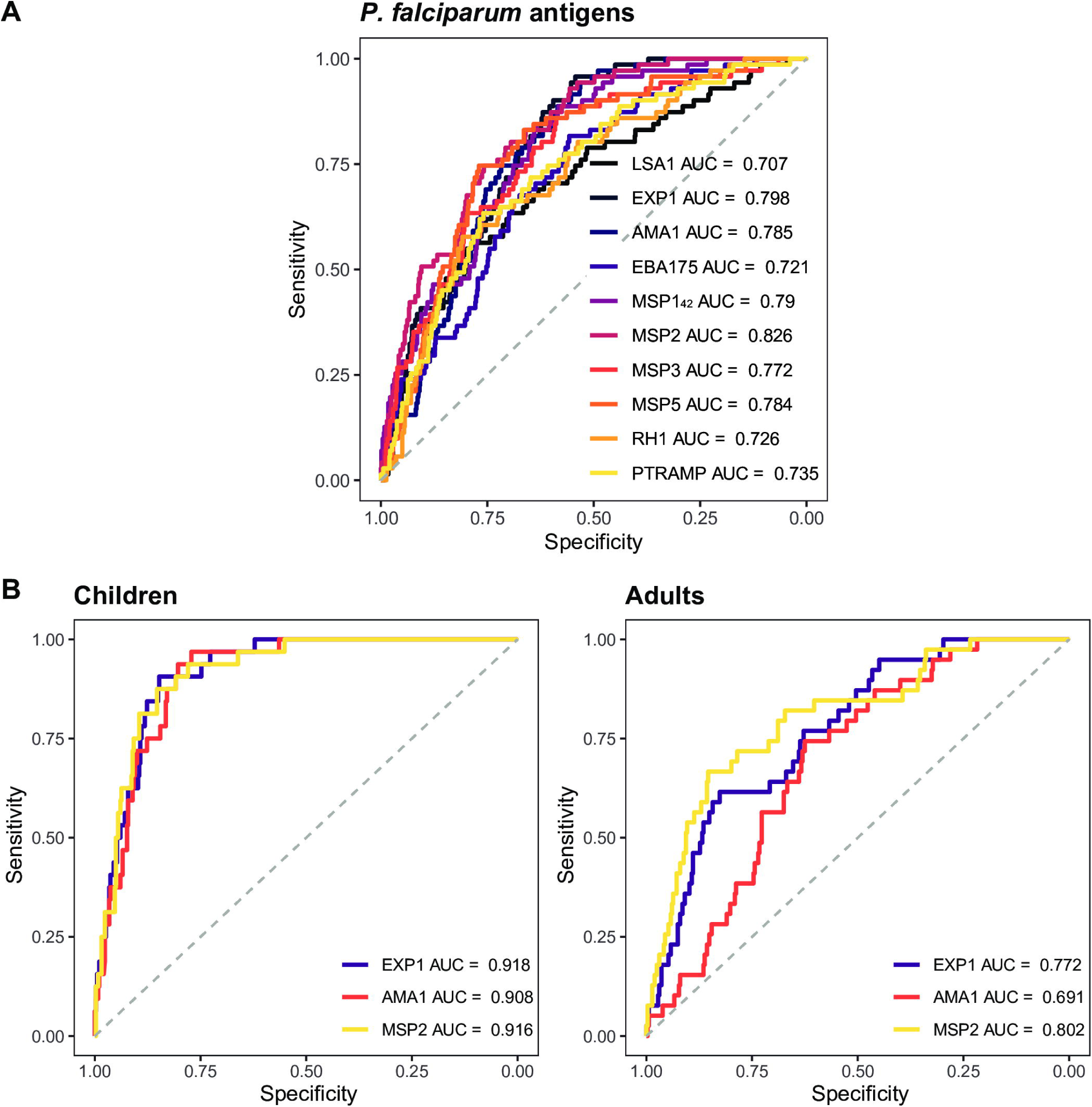
Receiver operating characteristic curves and their corresponding areas under the curve from the best performing IgG responses in detecting current *Plasmodium falciparum* infection. A. IgG responses (log_10_-transformed median fluorescence intensity [MFI] levels) against the LSA1, EXP1, AMA1, EBA175, MSP1_42_, MSP2, MSP3, MSP5, Rh1 and PTRAMP in serum samples from 715 endemic individuals and 50 Spanish donors were used as predictor variables and qPCR results as response variable. **B.** IgG responses (log_10_-transformed MFI levels) against EXP1, AMA1 and MSP2 in serum samples from children (N = 363) and adults (N = 352) were used as predictor variables and qPCR results were used as response variable. In each age group, antibody levels in samples from the 50 Spanish donors were used as negative controls. AUC: Area Under the Curve.

After stratifying by age group, AUC values improved for most of the antigens in children except for CelTOS, LSA1, MSP1 block 2 MAD20, RH1 and PTRAMP, for which the values were higher in adults (**Fig 1D** and **Table S2**). Remarkably, AUC values in children reached high levels (>0.9) for EXP1, AMA1 and MSP2 (**Fig 1D**, **Fig 5B** and **Table S2**). These top performers were used to calculate all possible combinations for the analysis in the children group, while CelTOS, LSA1, EXP1, MSP1 block 2, MSP1_42_, MSP2, MSP3, MSP5, RH1 and PTRAMP were used for the adult group. The best combinations and their corresponding AUCs were: EXP1 + AMA1 + MSP2 with AUC = 0.942 for children (**Fig S1C**) and LSA1 + EXP1 + MSP142 + MSP2 + MSP3 + MSP5 + PTRAMP with AUC = 0.848 for adults (**Fig S1D**). In addition, when all individuals with helminth infections were removed from the analysis, the AUC values slightly improved for most antigens with moderate AUC in the analysis including adults and children together (**Table S3**).

### Performance of antibodies as markers of exposure

Besides current infection, in some contexts it may be useful to be able to detect and quantify exposure. The cross-sectional design of this study did not allow for evaluating recent versus past exposure. Therefore, we explored the ability of serology to differentiate individuals from a non-endemic area (NC) from exposed individuals from an endemic area. To do so, we took as reference the EM cutoff, which splits the Mozambican individuals into two populations (seronegative and seropositive) based on the distribution of antibody levels to each antigen. We considered as good markers of exposure those antibodies that had a high specificity, i.e., those for which levels in the non-endemic population had little or no overlap with levels in the seropositive individuals from the endemic population, as defined by the EM cutoff.

Based on this, IgG levels against As37 (from *Ascaris* spp.), NIE (from *S. stercoralis*) and MEA (from *Schistosoma* spp.) were able to perfectly separate the non-endemic population from the seropositive endemic population (**Fig 6**). As16 (from *Ascaris* spp.), NaAPR1, NaGST1, NaSAA2 (from hookworm) and Tm16 (from *Trichuris* spp.) also had quite good performance in distinguishing the non-endemic population from the seropositive endemic population with the only exceptions of a few outliers among the non-exposed individuals (**Fig 6**). On the contrary, IgG levels against AyCp2 (from hookworm), TmWAP (from *Trichuris* spp.) and Sm25 (from *Schistosoma* spp.), were elevated in the non-endemic population (**Fig 6**) suggesting low specificity.

**Fig 6.**
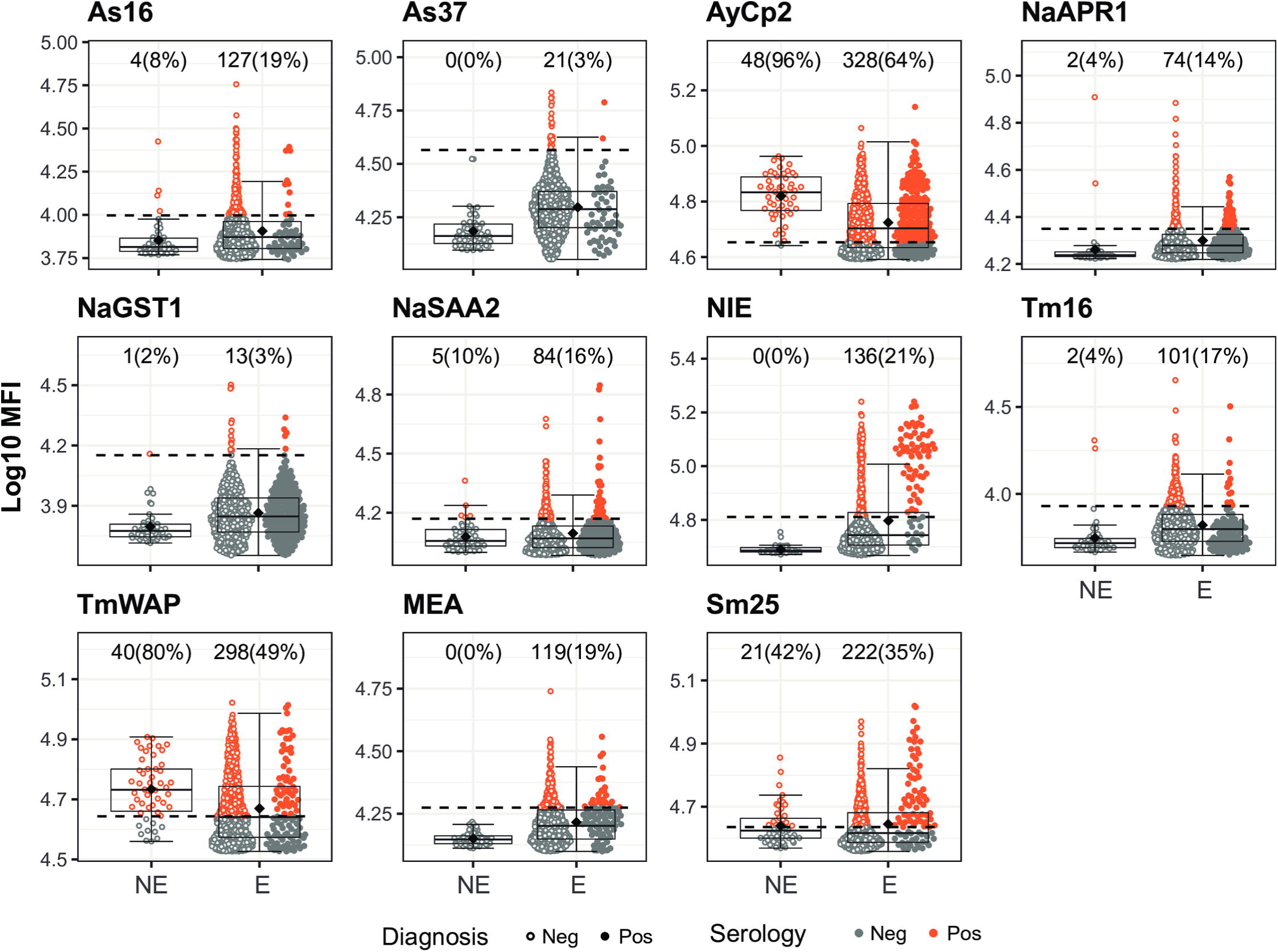
Helminth antigen-specific IgG levels in non-endemic and endemic populations and seropositivity cutoffs by the Expectation-Maximization algorithm. Antibody levels are presented as the log_10_-transformed median fluorescence intensity (MFI) of IgG against helminth antigens. In the endemic population, filled dots highlight individuals with a helminth infection with the corresponding parasite for each antigen detected by microscopy and/or qPCR. The dashed line represents the serology cutoff calculated with the Expectation-Maximization algorithm. Individuals above (orange) or below (grey) this cutoff were considered seropositive or seronegative, respectively. At the top of each plot, the absolute number and percentage of seropositive individuals with respect to the Non-Endemic (NE) (N = 50) or Endemic (E) (N = 715) populations are indicated. The boxplots represent the median (bold line), the mean (black diamond), the first and third quartiles (box) and the largest and smallest values within 1.5 times the interquartile range (whiskers). Data beyond the end of the whiskers are outliers.

In the case of *P. falciparum* antigens, IgG levels against α-gal, MSP1 block 2 and MSP2 perfectly distinguished the non-endemic population from the seropositive endemic population (**Fig 7**). IgG levels against AMA1, EBA175, EXP1, LSA1, MSP1_42_, MSP5, RH1, RH5 and PTRAMP also distinguished the non-endemic population from the seropositive endemic population with the only exceptions of a few outliers among the non-exposed individuals (**Fig 7**). Finally, IgG levels against CelTOS, MSP3, P41 and, specially, SSP2, were elevated in the non-endemic population (**Fig 7**).

**Fig 7.**
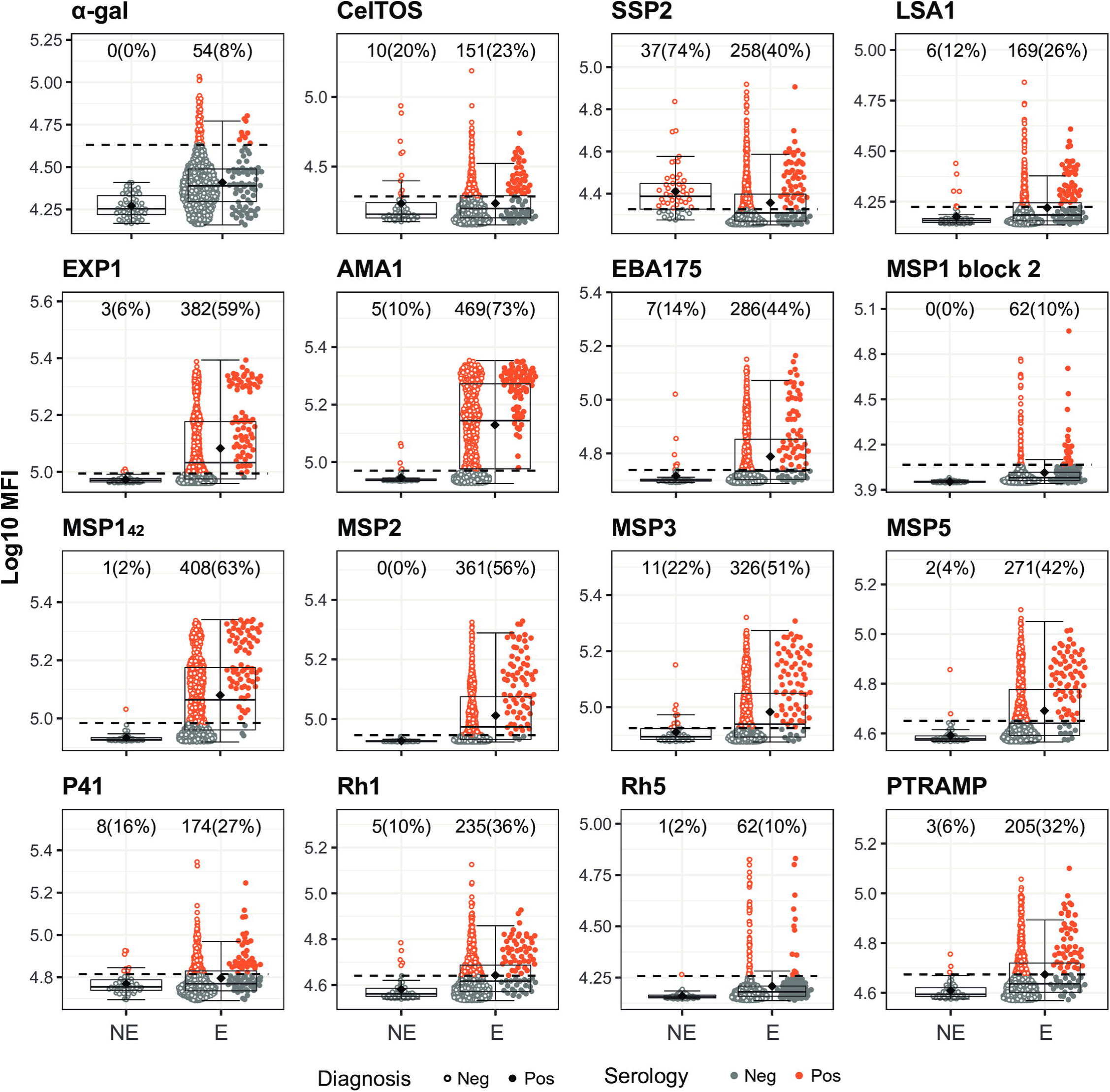
*Plasmodium falciparum* antigen-specific IgG levels in non-endemic and endemic populations and seropositivity cutoffs by the Expectation-Maximization algorithm. Antibody levels are presented as the log_10_-transformed median fluorescence intensity (MFI) of IgG against *P. falciparum* antigens. In the endemic population, filled dots highlight individuals with a *P. falciparum* infection detected by qPCR. The dashed line represents the serology cutoff calculated with the Expectation-Maximization algorithm. Individuals above (orange) or below (grey) this cutoff were considered seropositive or seronegative, respectively. At the top of each plot, the absolute number and percentage of seropositive individuals with respect to the Non-Endemic (NE) (N = 50) or Endemic (E) (N = 715) populations are indicated. The boxplots represent the median (bold line), the mean (black diamond), the first and third quartiles (box) and the largest and smallest values within 1.5 times the interquartile range (whiskers). Data beyond the end of the whiskers are outliers.

### Alternative seropositivity cutoffs for current infection

To further explore alternative ways to detect current infection, we investigated other possible seropositivity cutoffs extracted from the ROC analysis that could serve in different scenarios that would require prioritization of specificity, sensitivity or both. We also explored the ability of the EM cutoff to detect current infection. Finally, we assessed the performance of the classical cutoff based on the IgG levels measured in samples from a naïve population, in this case the Spanish donors, calculated as the mean plus 3 standard deviations. The results are summarized in **Fig 8** for the top performer antigens of the panel (NIE, MEA, Sm25, EXP1, AMA1 and MSP2) and for the rest of the antigens in **Fig S2**, **Fig S3** and **Table S4**, where the specificity and sensitivity of each cutoff method are indicated.

**Fig 8.**
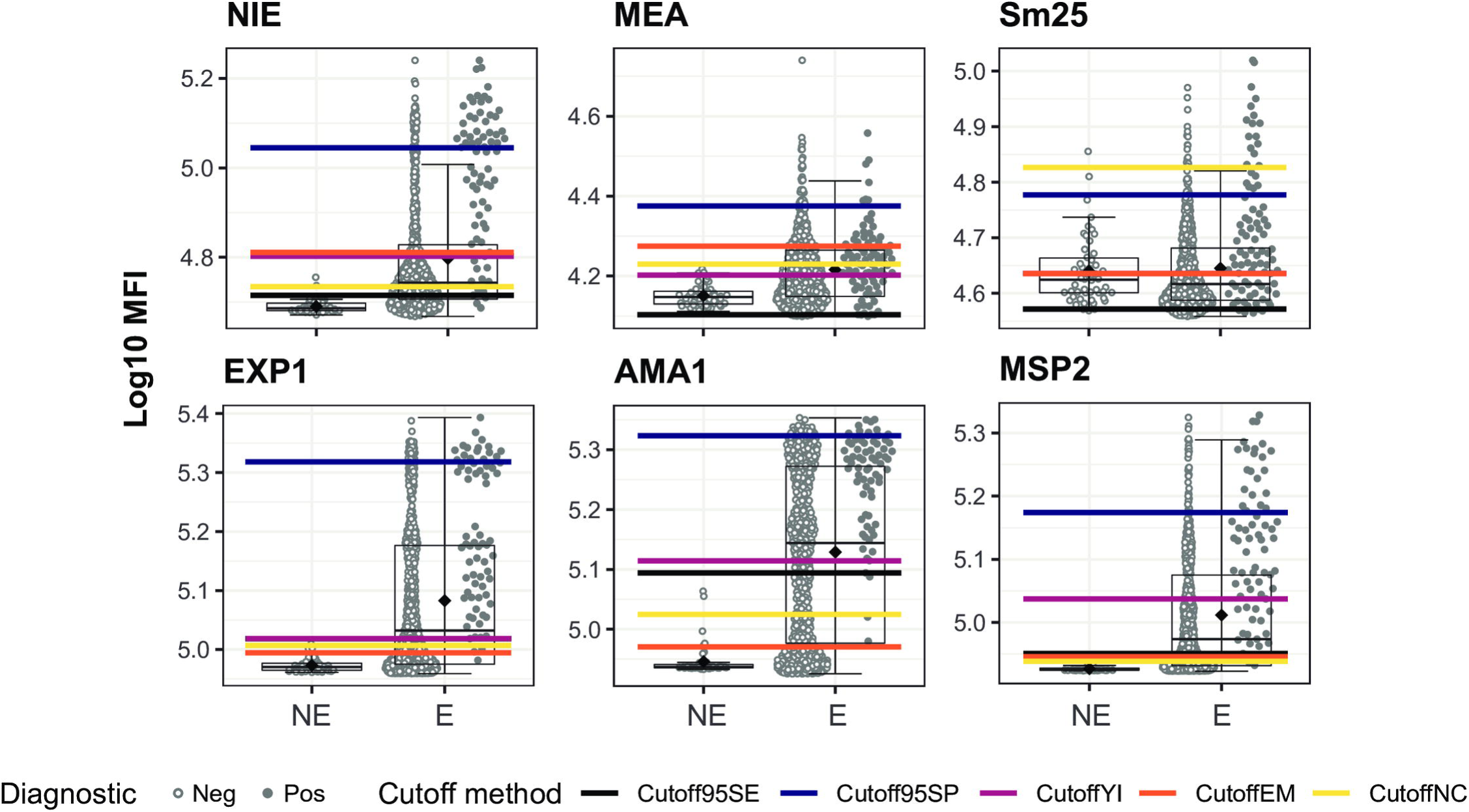
Helminth and *Plasmodium falciparum* antigen-specific IgG levels in non-endemic and endemic populations and seropositivity cutoffs calculated by different methods. Antibody levels are presented as the log_10_-transformed median fluorescence intensity (MFI) of IgG against *Strongyloides stercoralis* (NIE), *Schistosoma* spp. (Sm25 and MEA) and *P. falciparum* (EXP1, AMA1 and MSP2) antigens. In the endemic population, filled dots highlight individuals with a helminth infection with the corresponding parasite for each antigen detected by microscopy and/or qPCR in the case of helminths and qPCR in the case of *P. falciparum*. The boxplots represent the median (bold line), the mean (black diamond), the first and third quartiles (box) and the largest and smallest values within 1.5 times the interquartile range (whiskers). Data beyond the end of the whiskers are outliers. Cutoff95SE: cutoff prioritizing a minimum sensitivity of 95%; Cutoff95SP: cutoff prioritizing a minimum specificity of 95%; CutoffYI: cutoff corresponding to the maximum Youden’s Index; CutoffEM: cutoff defined by the Expectation-Maximization algorithm; CutoffNC: cutoff calculated as median + 3*standard deviations of the log_10_-transformed MFI levels from the non-endemic population. NE: Non-Endemic (N = 50), E: Endemic (N = 715). The rest of the antigens are in supplementary Fig S2 and S3.

For NIE specially, the EM cutoff provided a good trade-off between specificity (∼80 %) and sensitivity (∼80 %) and it overlapped with the Youden’s Index cutoff (**Fig 8** and **Table S4**). For Sm25, when taking *Schistosoma* spp. infection as a reference defined by the combination of microscopy and qPCR, the EM cutoff also overlapped with the Youden’s Index cutoff but the sensitivity was compromised (∼70%) by the large variability in the NC, as reflected by the elevated NC cutoff (**Fig 8** and **Table S4)**. On the other hand, responses against MEA induced less variability in the NC, but the EM and Youden’s Index cutoffs did not overlap. In this case, the EM cutoff had a 28% sensitivity and 82% specificity, while the Youden’s Index cutoff had a 70% sensitivity and 56% specificity (**Fig 8** and **Table S4**) when taking *Schistosoma* spp. infection as a reference defined by the combination of microscopy and qPCR.

For EXP1, AMA1 and MSP2, all cutoffs except the cutoff that prioritized specificity had sensitivity values above 94% and specificity values between 30-55% with the exception of the Youden’s Index cutoff for MSP2 (∼75% sensitivity and specificity) (**Fig 8** and **Table S4**).

### Effect of other infections on parasite-specific antibody levels

We assessed the effect of each single infection analyzed on the antigen-specific IgG levels of the top performer antigens of the panel (NIE, MEA, Sm25, EXP1, AMA1 and MSP2) to evaluate if other infections could be associated with false seropositive responses. *S. stercoralis* single infection was associated with statistically significantly higher IgG levels to MEA, Sm25, EXP1, AMA1 and MSP2 compared to no infections (**Fig 9**). In addition, *Schistosoma spp.* single infection was associated with statistically significantly higher IgG levels to NIE, AMA1 and MSP2 compared to no infections (**Fig 9**). Conversely, infection with *T. trichiura* was associated with statistically significantly lower IgG levels to MEA, EXP1, AMA1 and MSP2 compared to those with no infections (**Fig 9**). The same patterns for *S. stercoralis*, *Schistosoma* spp. and *T. trichiura* infections were observed among the rest of the antigens of the panel (**Fig S4** and **Fig S5**). We also evaluated how the proposed cutoffs would classify those individuals with increased antibody levels that have a negative diagnosis for the corresponding parasite to the antigen. Taking as an example NIE, only the 95% specificity cutoff classifies as seronegative most of those individuals that are *S. stercoralis* negative but *Schistosoma* spp. positive (**Fig 9**). However, this cutoff has a very reduced sensitivity (**Fig 9** and **Table S4**). This applies also to the rest of the top performing antigens, although in the case of Sm25, the NC cutoff has the highest specificity (**Fig 9** and **Table S4**).

**Fig 9.**
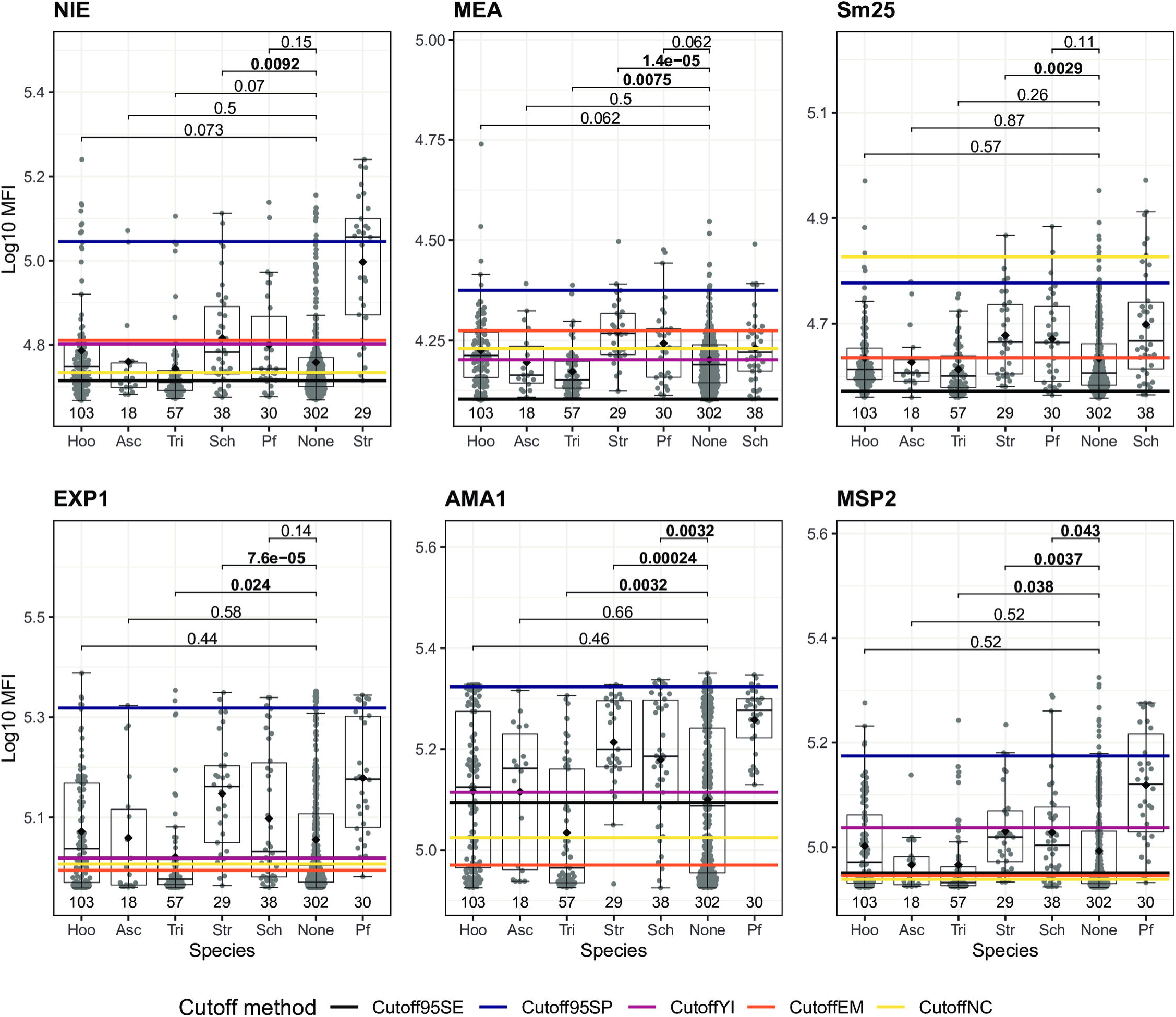
Effect of single infections on IgG levels to other infections. Only individuals with single infections or no infections at all detected by the combination of microscopy and qPCR are shown. Antibody levels are presented as the log_10_-transformed median fluorescence intensity (MFI) of IgG against *Strongyloides stercoralis* (NIE), *Schistosoma* spp. (Sm25 and MEA) and *P. falciparum* (EXP1, AMA1 and MSP2) antigens. For each antigen, antibody levels for each species were compared to the levels of those with no infection and the corresponding species to the antigen was shown as a reference. Statistical comparison between groups was performed by Wilcoxon rank-sum test and the adjusted P values by the Holm approach are shown. Statistically significant P values are highlighted in bold. The sample size of each group is shown below each boxplot. The boxplots represent the median (bold line), the mean (black diamond), the first and third quartiles (box) and the largest and smallest values within 1.5 times the interquartile range (whiskers). Data beyond the end of the whiskers are outliers. Cutoff95SE: cutoff prioritizing a minimum sensitivity of 95%; Cutoff95SP: cutoff prioritizing a minimum specificity of 95%; CutoffYI: cutoff corresponding to the maximum Youden’s Index; CutoffEM: cutoff defined by the Expectation-Maximization algorithm; CutoffNC: cutoff calculated as median + 3*standard deviations of the log_10_-transformed MFI levels from the non-endemic population. The rest of the antigens are in supplementary Fig S4 and S5.

## DISCUSSION

Antibody serology is a useful tool that can be exploited not only for understanding and detecting immunity to infections, but also for monitoring exposure and current infection. Here, we evaluated the ability of serology, measuring total IgE and specific IgG against a panel of STH, *Schistosoma* spp. and *P. falciparum* antigens, to detect current infection compared to microscopy, molecular diagnosis or the combination of both diagnosis, and to detect past exposure. Our study has proved the feasibility to use qSAT to measure antibody levels against a large panel of parasite antigens in serum eluted from DBS. The fact that Luminex allows for multiplexing several antigens with a very small amount of sample shows an advantage over conventional ELISAs, since it could be easily implemented for the detection of antibody responses to several pathogens simultaneously, and allows the integration of control programs for different pathogens. In addition, finger-pricking for the collection of DBS is a minimally invasive technique and is particularly practical for remote health and surveillance programs, since it offers simplified logistics and it is more cost effective than serum collection.

While IgG levels against most of the helminth antigens included in our panel were not good markers of current infection, many seem to be good enough markers of exposure. IgG to NIE (*S. stercoralis*), MEA and Sm25 (*Schistosoma* spp.), and MSP2 (*P. falciparum*) antigens showed the most promising results in detecting current infection. This is of particular interest for *S. stercoralis* due to the need for improved diagnosis. Microscopic examination has a very low sensitivity (39, 40), as we have also verified by comparing the low number of *S. stercoralis* positive individuals detected by microscopy with those detected by qPCR. However, qPCR and other methods with better sensitivity than microscopy, such as the Baermann technique and Koga agar plate culture, could still leave out some infections given the frequency of low-burden infections (40). Serological tests based on the detection of antibodies to whole *S. stercoralis* parasites or crude extracts have already shown excellent results (39, 40). Nonetheless, the need for a constant supply of parasites and variability of the source are limitations of those assays (41). On the other hand, antibody serological methods based on recombinant proteins, such as NIE (42), are more reproducible. Recently, a study evaluated the performance of two ELISAs that detect IgG and IgG4 against the NIE and SsRI antigens and found specificities between 91-98% and sensitivities between 78-92% (43). In our study, the cutoff with the maximum Youden’s index results in specificity and sensitivity of around 79% and 85%, respectively. One source of variations in accuracy is cross-reactivity. For example, the AUC for IgG against NIE increased and reached high accuracy after elimination of those samples from participants with other helminth and *P. falciparum* infections detected in our study. This informs of a possible cross-reactivity with other species and therefore accuracy might vary depending on the prevalence of other infections in the target area. This is particularly relevant for international travelers and people living in very low prevalence settings for co-infections. However, a study using NIE serology found no cross-reactivity between *S. stercoralis* and infections with other STH (44).

Interestingly, our results indicate that infection by *Schistosoma* spp. was associated with increased levels to NIE, AMA1 and MSP2, among others in the panel, and infection by *S. stercoralis* was associated with elevated IgG levels to all antigens. Such generalized pattern would suggest other mechanisms rather than cross-reactivity, like induction of polyreactive antibodies (45, 46). Another possibility is that increased antibody levels simply reflect an increased susceptibility and exposure to infections among *Schistosoma* spp. and *S. stercoralis* infected individuals. In any case, it should be considered that these infections might reduce the specificity of the assay and these results underscore the importance to evaluate antibody serological tools in areas of co-endemicity. Conversely, the opposite pattern has been associated with *T. trichiura* infected individuals, in which IgG levels were generally lower than the non-infected individuals. This suggests a possible immunomodulation by *T. trichiura* as observed by others (47) which could reduce the sensitivity of the assay.

The WHO included the control of morbidity caused by *S. stercoralis* as one of its objectives for 2030 (1) and in September 2020 there was a WHO meeting to discuss diagnostic methods for the control of strongyloidiasis (11). The control strategy is based on the distribution of ivermectin using established platforms to control other STH, and pilot interventions are currently being evaluated (1). Such control strategies require monitoring of the transmission in order to guide future steps of the program. According to the meeting, serological assessment by NIE ELISA is the best available option, although it is not perfect and it should be accompanied by a Baermann or agar-plate method whenever possible (11). NIE has been used in different platforms including ELISA, Luminex and luciferase immunoprecipitation system (LIPS) (43,44,48). Also, it has been proposed as a valuable point-of-care diagnostic tool for its use in lateral flow cassettes (49, 50). While NIE serology shows promising results as a diagnostic tool, it is important to consider that total serological reversion is unlikely to occur and, therefore, differentiating between past exposure and current infection can be challenging. However, unlike for other parasitic infections, *S. stercoralis*-specific antibodies decrease markedly within the following months post-treatment (11, 12) and, as we and others have demonstrated, it is possible to achieve a moderate to high accuracy based on antibody levels. This is particularly useful to evaluate the success of chemotherapy or MDA and verify transmission interruption (14, 15).

Regarding *Schistosoma* spp., antibody serology is a valuable screening tool in non-endemic or low-transmission areas (19). In addition, the WHO strategy for control of *Schistosoma* spp. infections is based on administration of preventive chemotherapy at large scale. Therefore, for the same reasons mentioned above for *S. stercoralis*, serology could be a valuable tool to inform decisions in the program. In our study, the cutoff with the maximum Youden’s index for IgG against MEA and Sm25 antigens showed sensitivities of 59-84% and 56-75%, respectively, and specificities of 54-69% and 63-85%, respectively. The wide range is affected by the diagnostic method and the species used as a reference. In fact, the genus-specific qPCR was a potential limitation since *S. haematobium* can also be detected in feces (51). Therefore, the inability to distinguish species could have affected the accuracy of MEA and Sm25. In addition, better specificities and sensitivities are also possible depending on the cutoff method selected. Other studies with other antigens and platforms have found also a wide range of specificity and sensitivity with different assays (18, 19) and have reported a limited utility of *Schistosoma* spp. serology in the detection of current infection in individuals from endemic areas or in the follow-up after treatment (41). The reason for this is that antibody levels may remain elevated for a long period of time after elimination of the infection, contrary to what it is observed for *S. stercoralis* (19, 52). The Sm25 antigen has been evaluated previously and it was proposed as an excellent antigen for measuring responses to *S. mansoni* (53). However, we observed a similar accuracy when the reference species was *S. haematobium*. Thus, it most likely detects both *Schistosoma* species due to cross-reactivity.

Specific IgG against many of the *P. falciparum* antigens from our panel were moderately good in detecting current infection, especially IgG against MSP2. In addition, for EXP1, AMA1 and MSP2, the accuracy particularly increased to high levels in children after stratification by age group, presumably because there is less interference from past exposures compared to adults, as observed also by others for other *P. falciparum* antigens (27). IgG levels against these antigens were also very low in the non-endemic population. Given the long-lived nature of antibody responses, especially in endemic areas, it is unlikely that antibody responses find a niche among *P. falciparum* diagnostic methods, although others have identified antibody responses against different antigens from *P. falciparum* (54, 55) and *P. vivax* (56) as potential markers of recent exposure. Our results suggest that IgG against the antigens in our panel could be useful markers in the context of control and elimination programs to monitor transmission using children as sentinels in medium-to high-transmission areas, where long-lasting antibody responses in adults might not represent the current transmission scenario. In addition, serology might be particularly relevant in areas of low transmission where large sample size are required to monitor prevalence and the use of other methods such as qPCR or microscopy would require high costs (27), time and would not be able to capture recent exposure. Additional longitudinal studies with a larger sample size in different age groups and areas of varying transmissions would be necessary to corroborate the utility of the markers proposed here as reliable indicators of infection. Interestingly, our results align with others that found IgG to MSP2 among the top 10 responses predictive of malaria incidence in study participants (57) and a strong correlation of IgG to AMA1 with both clinical malaria and asymptomatic infection (55). However, the use of these antigens also comes with potential disadvantages. MSP2 is a highly polymorphic antigen (58, 59) and therefore the genotypes prevalent in the community might differ from that included in the assay (60). As for EXP1, and specially AMA1, their high immunogenicity leads to high levels that reach saturation since early age at high transmission intensities, but they might be suitable for low transmission intensities (60). Interestingly, the FlexMAP 3D platform from Luminex allowed has a wider dynamic range compared to other platforms such as the Luminex 200 or ELISA, probably allowing for the detection of increases induced by current infections with respect to the basal levels even for highly immunogenic antigens.

The comparison of seropositivity cutoffs estimated by different methods highlights the importance of setting the cutoffs based on the antibody data from the endemic target population, as opposed to the classic approach of using data from a non-endemic population as reference. The reason behind is to take background responses into account, which might differ depending on the origin of the samples. For instance, we have observed here an elevated NC cutoff for some antigens, which is suggestive of the presence of antibodies in the non-endemic population that cross-react with some of the antigens in our panel. We have provided an approach to calculate the seropositivity cutoffs based on the distribution of antibody levels in an endemic population using the EM algorithm, also previously used by others (61). Other cutoffs extracted from the ROC curves provided varying sensitivities and specificities, sometimes at the expense of a much reduced balance between these parameters, which can be used in different scenarios. For example, in the context of public health, in low prevalence settings or control programs, particularly towards the final elimination stage, high sensitivity with a prioritization of a high specificity is desirable since it reduces the number of false positive results and saves unnecessary interventions (62). On the contrary, in the context of individual-focused diagnosis, sensitivity might be prioritized over specificity. In non-endemic areas where the number of imported cases is low, it may be preferable to treat a false positive patient than leaving out a potential infection that can be more threatening in people from non-endemic settings due to inexistent immunity.

In addition to IgG, IgG subclasses or other isotypes could be more useful for different species given the properties of each antibody type. For example, here we evaluated also total IgE responses, which are a well-known hallmark of the Th2 predominant responses typical of helminth infections (63). IgE has a shorter half-life than IgG (∼3 days versus ∼21 days), which makes it a good candidate of recent exposure to helminths (64). We have found that total IgE levels are a good marker of hookworm and *Schistosoma spp.* recent infections with moderate accuracy. Besides IgE, other studies have suggested *Ascaris*-specific IgG4 to perform better since it showed less cross-reactivity with other helminths than IgG1, IgG2 and IgG3 (65, 66). Moreover, for *P. falciparum*, the long-standing issue of finding a good marker of recent exposure could also benefit from combining the right antigen with the detection by IgG3 antibodies. IgG3 is one of the main IgG subclasses produced during *Plasmodium* spp. infection and it has a shorter half-life compared to the rest of IgG subclasses (∼7 days versus ∼21 days) (67).

Many of the antigens included in the analysis were not reliable markers of current infection since they were not able to classify correctly the population based in the reference diagnostic methods and reach an AUC > 0.7. On one hand, IgG levels may stay elevated long after acute infection, leading to high residual responses from past infections in the exposed non-infected population. On the other hand, due to antibody dynamics, there is a window of time between infection and detection of antibody titers that should also be taken into account. For example, in our study, all malaria cases included were asymptomatic and it could be that many of these infections were recent. In addition, the specificity of the responses might be against antigens from a specific stage of the life cycle of the parasite that has not been reached yet. Other possible reasons for lack of accuracy are: i) the lack of a proper gold standard diagnostic method for some parasites; ii) the low immunogenicity of some antigens; iii) and shared epitopes between antigens leading to cross-reactive antibodies in the non-infected population. In relation to the lack of a proper gold standard for comparison, serology might be able to capture infections that yield a low number of diagnostic forms for microscopy such as chronic *Schistosoma* spp. infections, which are characterized by low o absent egg production (68). For this reason, conventional microscopy is not completely optimal for the diagnosis of *Schistosoma* spp. and using it as a reference may result in a false low sensitivity.

The design of the current study is subject to other limitations. The Telemann technique used in this study for the detection of *S. stercoralis* by microscopy is not among the recommended methods and therefore we consider the finding of *S. stercoralis* by microscopy purely anecdotic. Moreover, although qPCR has in general a better sensitivity than microscopy, it should be noted that it can also detect genetic material of dead parasites after resolving the infection. In addition, it would have been ideal to have longitudinal data to properly evaluate exposure and time since infection within the endemic population. We also acknowledge the difficulty of field implementation of Luminex equipment in low- and middle-income settings, but a reference lab with the necessary equipment could be in charge of processing the samples. Finally, for *P. falciparum* analysis, the participants of our study were all asymptomatic, which can be a limitation since antibody signatures might differ compared to symptomatic (69). However, this also shows the ability of antibody serology against specific antigens to detect an active infection even at low density.

In conclusion, we provide evidence for the utility of serology as a marker of current infection with some parasitic infections of public health importance in a co-endemic population from Southern Mozambique. We corroborate the reported good accuracy of IgG against the NIE (*S. stercoralis)* and show that IgG against MEA, Sm25 (*Schistosoma* spp.), EXP1, AMA1 and MSP2 (*P. falciparum*) might also be markers of current infection reliable enough to be employed as decision-making tools within control and elimination programs. Importantly, antibody serology by Luminex allows for multiplexing and, therefore, integration of control programs for different pathogens in co-endemic areas. Furthermore, we also show that IgG seropositivity against many of the antigens differentiated the seropositive endemic population from the non-endemic populations, suggesting a possible role as markers of exposure.

## Data Availability

All data produced in the present study are available upon reasonable request to the authors

## ACKNOWLEDGEMENTS

We are grateful to the volunteers and their families and to all field workers and lab technicians that participated in this study. We also thank CISM Demography and Social Sciences department for their indispensable assistance during field work. Special thanks to the team members from ISGlobal, specifically Laura Puyol for the organization and coordination of shipment of samples, Pau Cisteró for the assistance in setting up the *P. falciparum* qPCR, Diana Barrios and Alfons Jiménez for the technical support in the lab, Miquel Vázquez-Santiago and Llorenç Quintó for the statistical advice, and Chenjerai Jairoce for the aid in assay optimization. For antigen procurement, we also thank Thomas Nutman (NIH/NIAID, USA), Sukwan Handali (CDC, USA) and Deepak Gaur (ICGEB and Jawaharlal Nehru University, India). On qPCR analysis, we particularly would like to thank Eric Brienen (LUMC, the Netherlands).

## REFERENCES

1. World Health Organization. Soil-transmitted helminth infections [Internet]. 2020. Available from: https://www.who.int/news-room/fact-sheets/detail/soil-transmitted-helminth-infections

2. World Health Organization. Schistosomiasis [Internet]. 2021. Available from: https://www.who.int/news-room/fact-sheets/detail/schistosomiasis

3. World Health Organization. World malaria report 2021. Geneva; 2021.

4. Salgame P, Yap GS, Gause WC. Effect of helminth-induced immunity on infections with microbial pathogens. Nat Immunol. 2013 Nov;14(11):1118–26.

5. Doehring E, Feldmeier H, Daffalla AA. Day-to-day variation and circadian rhythm of egg excretion in urinary schistosomiasis in the Sudan. Ann Trop Med Parasitol. 1983 Dec;77(6):587–94.

6. Hall A. Quantitative variability of nematode egg counts in faeces: a study among rural Kenyans. Trans R Soc Trop Med Hyg. 1981;75(5):682–7.

7. Anderson RM, Schad GA. Hookworm burdens and faecal egg counts: an analysis of the biological basis of variation. Trans R Soc Trop Med Hyg. 1985;79(6):812–25.

8. Dreyer G, Fernandes-Silva E, Alves S, Rocha A, Albuquerque R, Addiss D. Patterns of detection of Strongyloides stercoralis in stool specimens: implications for diagnosis and clinical trials. J Clin Microbiol. 1996 Oct;34(10):2569–71.

9. Werkman M, Wright JE, Truscott JE, Oswald WE, Halliday KE, Papaiakovou M, et al. The impact of community-wide, mass drug administration on aggregation of soil-transmitted helminth infection in human host populations. Parasit Vectors. 2020;13(1):290.

10. Gitta B, Kilian N. Diagnosis of Malaria Parasites Plasmodium spp. in Endemic Areas: Current Strategies for an Ancient Disease. BioEssays. 2020 Jan 1;42(1):1900138.

11. World Health Organization. Diagnostic methods for the control of strongyloidiasis, Virtual meeting, 29 September 2020. Geneva; 2020.

12. Buonfrate D, Sequi M, Mejia R, Cimino RO, Krolewiecki AJ, Albonico M, et al. Accuracy of Five Serologic Tests for the Follow up of Strongyloides stercoralis Infection. PLoS Negl Trop Dis. 2015 Feb 10;9(2):e0003491.

13. Balachandra D, Ahmad H, Arifin N, Noordin R. Direct detection of Strongyloides infection via molecular and antigen detection methods. Eur J Clin Microbiol Infect Dis Off Publ Eur Soc Clin Microbiol. 2021 Jan;40(1):27–37.

14. Marks M, Gwyn S, Toloka H, Kositz C, Asugeni J, Asugeni R, et al. Impact of Community Treatment With Ivermectin for the Control of Scabies on the Prevalence of Antibodies to Strongyloides stercoralis in Children. Clin Infect Dis [Internet]. 2020 Dec 15;71(12):3226–8. Available from: https://doi.org/10.1093/cid/ciaa584

15. Mounsey K, Kearns T, Rampton M, Llewellyn S, King M, Holt D, et al. Use of dried blood spots to define antibody response to the Strongyloides stercoralis recombinant antigen NIE. Acta Trop. 2014 Oct;138:78–82.

16. Vlaminck J, Supali T, Geldhof P, Hokke CH, Fischer PU, Weil GJ. Community Rates of IgG4 Antibodies to Ascaris Haemoglobin Reflect Changes in Community Egg Loads Following Mass Drug Administration. PLoS Negl Trop Dis. 2016 Mar;10(3):e0004532.

17. Logan J, Pearson MS, Manda SS, Choi Y-J, Field M, Eichenberger RM, et al. Comprehensive analysis of the secreted proteome of adult Necator americanus hookworms. PLoS Negl Trop Dis. 2020 May 26;14(5):e0008237.

18. Kinkel H-F, Dittrich S, Bäumer B, Weitzel T. Evaluation of eight serological tests for diagnosis of imported schistosomiasis. Clin Vaccine Immunol. 2012/03/21. 2012 Jun;19(6):948–53.

19. Hinz R, Schwarz NG, Hahn A, Frickmann H. Serological approaches for the diagnosis of schistosomiasis – A review. Mol Cell Probes. 2017;31:2–21.

20. Drakeley C, Cook J. Chapter 5 Potential Contribution of Sero-Epidemiological Analysis for Monitoring Malaria Control and Elimination: Historical and Current Perspectives. In Academic Press; 2009. p. 299–352.

21. Drakeley CJ, Corran PH, Coleman PG, Tongren JE, McDonald SLR, Carneiro I, et al. Estimating medium- and long-term trends in malaria transmission by using serological markers of malaria exposure. Proc Natl Acad Sci U S A [Internet]. 2005 Apr 5;102(14):5108 LP–5113. Available from: http://www.pnas.org/content/102/14/5108.abstract

22. Won KY, Sambou S, Barry A, Robinson K, Jaye M, Sanneh B, et al. Use of Antibody Tools to Provide Serologic Evidence of Elimination of Lymphatic Filariasis in The Gambia. Am J Trop Med Hyg. 2018;98(1):15–20.

23. Guevara Á, Salazar E, Vicuña Y, Hassan HK, Muro A, Guderian R, et al. Use of Ov16-Based Serology for Post-Elimination Surveillance of Onchocerciasis in Ecuador. Am J Trop Med Hyg. 2020;103(4):1569–71.

24. Pinsent A, Solomon AW, Bailey RL, Bid R, Cama A, Dean D, et al. The utility of serology for elimination surveillance of trachoma. Nat Commun. 2018;9(1):5444.

25. Rabello AL, Garcia MM, Pinto da Silva RA, Rocha RS, Katz N. Humoral immune responses in patients with acute Schistosoma mansoni infection who were followed up for two years after treatment. Clin Infect Dis an Off Publ Infect Dis Soc Am. 1997 Mar;24(3):304–8.

26. Doenhoff MJ, Chiodini PL, Hamilton J V. Specific and sensitive diagnosis of schistosome infection: can it be done with antibodies? Trends Parasitol. 2004 Jan;20(1):35–9.

27. Markwalter CF, Nyunt MH, Han ZY, Henao R, Jain A, Taghavian O, et al. Antibody signatures of asymptomatic Plasmodium falciparum malaria infections measured from dried blood spots. Malar J. 2021;20(1):378.

28. Santano R, Rubio R, Grau-Pujol B, Escola V, Muchisse O, Cuamba I, et al. Plasmodium falciparum and Helminth Coinfections Increase IgE and Parasite-Specific IgG Responses. Microbiol Spectr. 2021 Dec 8;e0110921.

29. Saúte F, Aponte J, Almeda J, Ascaso C, Vaz N, Dgedge M, et al. Malaria in southern Mozambique: incidence of clinical malaria in children living in a rural community in Manhiça district. Trans R Soc Trop Med Hyg. 2003;97(6):655–60.

30. Grau-Pujol B, Martí-Soler H, Escola V, Demontis M, Jamine JC, Gandasegui J, et al. Towards soil-transmitted helminths transmission interruption: The impact of diagnostic tools on infection prediction in a low intensity setting in Southern Mozambique. Beechler BR, editor. PLoS Negl Trop Dis. 2021 Oct 25;15(10):e0009803.

31. World Health Organization. Basic laboratory methods in medical parasitology [Internet]. 1991. Available from: https://apps.who.int/iris/handle/10665/40793

32. Telemann W. Eine Methode zur Erleichterung der Auffindung von Parasiteneiern in den Faeces. Dtsch Medizinische Wochenschrift. 1908;34(35):1510–1.

33. Robin X, Turck N, Hainard A, Tiberti N, Lisacek F, Sanchez J-C, et al. pROC: an open-source package for R and S+ to analyze and compare ROC curves. BMC Bioinformatics. 2011 Mar 17;12:77.

34. Scrucca L, Fop M, Murphy TB, Raftery AE. mclust 5: Clustering, Classification and Density Estimation Using Gaussian Finite Mixture Models. R J. 2016 Aug;8(1):289–317.

35. Wickham H, Averick M, Bryan J, Chang W, McGowan L, François R, et al. Welcome to the Tidyverse. J Open Source Softw [Internet]. 2019 Nov 21 [cited 2021 Apr 20];4(43):1686. Available from: https://joss.theoj.org/papers/10.21105/joss.01686

36. Kassambara A. “ggplot2” Based Publication Ready Plots [R package ggpubr version 0.4.0]. 2020 Jun 27 [cited 2021 Apr 20]; Available from: https://cran.r-project.org/package=ggpubr

37. Clarke E, Sherrill-Mix SA. ggbeeswarm: Categorical scatter (violin point) plots [Internet]. R package version 0.6.0. 2017 [cited 2021 Apr 20]. Available from: https://rdrr.io/cran/ggbeeswarm/

38. Swets JA. Measuring the accuracy of diagnostic systems. Science. 1988 Jun;240(4857):1285–93.

39. Requena-Méndez A, Chiodini P, Bisoffi Z, Buonfrate D, Gotuzzo E, Muñoz J. The laboratory diagnosis and follow up of strongyloidiasis: a systematic review. PLoS Negl Trop Dis. 2013/01/17. 2013;7(1):e2002–e2002.

40. Buonfrate D, Formenti F, Perandin F, Bisoffi Z. Novel approaches to the diagnosis of Strongyloides stercoralis infection. Clin Microbiol Infect. 2015 Jun 1;21(6):543–52.

41. Tamarozzi F, Ursini T, Hoekstra PT, Silva R, Costa C, Gobbi F, et al. Evaluation of microscopy, serology, circulating anodic antigen (CAA), and eosinophil counts for the follow-up of migrants with chronic schistosomiasis: a prospective cohort study. Parasit Vectors. 2021 Mar 9;14(1):149.

42. Ravi V, Ramachandran S, Thompson RW, Andersen JF, Neva FA. Characterization of a recombinant immunodiagnostic antigen (NIE) from Strongyloides stercoralis L3-stage larvae. Mol Biochem Parasitol. 2002;125(1–2):73–81.

43. Tamarozzi F, Longoni SS, Mazzi C, Pettene S, Montresor A, Mahanty S, et al. Diagnostic accuracy of a novel enzyme-linked immunoassay for the detection of IgG and IgG4 against Strongyloides stercoralis based on the recombinant antigens NIE/SsIR. Parasit Vectors. 2021;14(1):412.

44. Krolewiecki AJ, Ramanathan R, Fink V, McAuliffe I, Cajal SP, Won K, et al. Improved diagnosis of Strongyloides stercoralis using recombinant antigen-based serologies in a community-wide study in northern Argentina. Clin Vaccine Immunol. 2010 Oct;17(10):1624–30.

45. Dimitrov JD, Planchais C, Roumenina LT, Vassilev TL, Kaveri S V, Lacroix-Desmazes S. Antibody Polyreactivity in Health and Disease: Statu Variabilis. J Immunol [Internet]. 2013 Aug 1;191(3):993 LP–999. Available from: http://www.jimmunol.org/content/191/3/993.abstract

46. Patel P, Kearney JF. Immunological Outcomes of Antibody Binding to Glycans Shared between Microorganisms and Mammals. J Immunol [Internet]. 2016 Dec 1;197(11):4201 LP–4209. Available from: http://www.jimmunol.org/content/197/11/4201.abstract

47. Esen M, Mordmüller B, de Salazar PM, Adegnika AA, Agnandji ST, Schaumburg F, et al. Reduced antibody responses against Plasmodium falciparum vaccine candidate antigens in the presence of Trichuris trichiura. Vaccine. 2012 Dec;30(52):7621–4.

48. Rascoe LN, Price C, Shin SH, McAuliffe I, Priest JW, Handali S. Development of Ss-NIE-1 Recombinant Antigen Based Assays for Immunodiagnosis of Strongyloidiasis. PLoS Negl Trop Dis. 2015;9(4):1–10.

49. Yunus MH, Arifin N, Balachandra D, Anuar NS, Noordin R. Lateral Flow Dipstick Test for Serodiagnosis of Strongyloidiasis. Am J Trop Med Hyg. 2019 Aug;101(2):432–5.

50. Noordin R, Osman E, Kalantari N, Anuar NS, Gorgani-Firouzjaee T, Sithithaworn P, et al. A point-of-care cassette test for detection of Strongyloides stercoralis. Acta Trop [Internet]. 2022;226:106251. Available from: https://www.sciencedirect.com/science/article/pii/S0001706X21004290

51. Cunin P, Tchuem Tchuenté L-A, Poste B, Djibrilla K, Martin PM V. Interactions between Schistosoma haematobium and Schistosoma mansoni in humans in north Cameroon. Trop Med Int Health. 2003 Dec;8(12):1110–7.

52. Yong MK, Beckett CL, Leder K, Biggs BA, Torresi J, O’Brien DP. Long-Term Follow-Up of Schistosomiasis Serology Post-Treatment in Australian Travelers and Immigrants. J Travel Med. 2010 Mar 1;17(2):89–93.

53. Njenga SM, Kanyi HM, Arnold BF, Matendechero SH, Onsongo JK, Won KY, et al. Integrated Cross-Sectional Multiplex Serosurveillance of IgG Antibody Responses to Parasitic Diseases and Vaccines in Coastal Kenya. Am J Trop Med Hyg. 2020 Jan;102(1):164–76.

54. van den Hoogen LL, Walk J, Oulton T, Reuling IJ, Reiling L, Beeson JG, et al. Antibody Responses to Antigenic Targets of Recent Exposure Are Associated With Low-Density Parasitemia in Controlled Human Plasmodium falciparum Infections [Internet]. Vol. 9, Frontiers in Microbiology. 2019. p. 3300. Available from: https://www.frontiersin.org/article/10.3389/fmicb.2018.03300

55. Wu L, Mwesigwa J, Affara M, Bah M, Correa S, Hall T, et al. Antibody responses to a suite of novel serological markers for malaria surveillance demonstrate strong correlation with clinical and parasitological infection across seasons and transmission settings in The Gambia. BMC Med. 2020;18(1):304.

56. Longley RJ, White MT, Takashima E, Brewster J, Morita M, Harbers M, et al. Development and validation of serological markers for detecting recent Plasmodium vivax infection. Nat Med [Internet]. 2020;26(5):741–9. Available from: https://doi.org/10.1038/s41591-020-0841-4

57. Helb DA, Tetteh KKA, Felgner PL, Skinner J, Hubbard A, Arinaitwe E, et al. Novel serologic biomarkers provide accurate estimates of recent Plasmodium falciparum exposure for individuals and communities. Proc Natl Acad Sci. 2015 Aug 11;112(32):E4438 LP–E4447.

58. Franks S, Baton L, Tetteh K, Tongren E, Dewin D, Akanmori BD, et al. Genetic diversity and antigenic polymorphism in Plasmodium falciparum: extensive serological cross-reactivity between allelic variants of merozoite surface protein 2. Infect Immun. 2003 Jun;71(6):3485–95.

59. Hoffmann EH, da Silveira LA, Tonhosolo R, Pereira FJ, Ribeiro WL, Tonon AP, et al. Geographical patterns of allelic diversity in the Plasmodium falciparum malaria-vaccine candidate, merozoite surface protein-2. Ann Trop Med Parasitol. 2001 Mar;95(2):117–32.

60. Corran P, Coleman P, Riley E, Drakeley C. Serology: a robust indicator of malaria transmission intensity? Trends Parasitol. 2007 Dec;23(12):575–82.

61. Migchelsen SJ, Martin DL, Southisombath K, Turyaguma P, Heggen A, Rubangakene PP, et al. Defining Seropositivity Thresholds for Use in Trachoma Elimination Studies. PLoS Negl Trop Dis. 2017 Jan 18;11(1):e0005230–e0005230.

62. Gass K. Time for a diagnostic sea-change: Rethinking neglected tropical disease diagnostics to achieve elimination. PLoS Negl Trop Dis. 2020 Dec 31;14(12):e0008933– e0008933.

63. Maizels RM, Balic A, Gomez-Escobar N, Nair M, Taylor MD, Allen JE. Helminth parasites – masters of regulation. Immunol Rev. 2004 Oct 1;201(1):89–116.

64. Lawrence MG, Woodfolk JA, Schuyler AJ, Stillman LC, Chapman MD, Platts-Mills TAE. Half-life of IgE in serum and skin: Consequences for anti-IgE therapy in patients with allergic disease. J Allergy Clin Immunol. 2016/07/14. 2017 Feb;139(2):422–428.e4.

65. Chatterjee BP, Santra A, Karmakar PR, Mazumder DN. Evaluation of IgG4 response in ascariasis by ELISA for serodiagnosis. Trop Med Int Health. 1996 Oct;1(5):633–9.

66. Santra A, Bhattacharya T, Chowdhury A, Ghosh A, Ghosh N, Chatterjee BP, et al. Serodiagnosis of ascariasis with specific IgG4 antibody and its use in an epidemiological study. Trans R Soc Trop Med Hyg. 2001;95(3):289–92.

67. Vidarsson G, Dekkers G, Rispens T. IgG subclasses and allotypes: From structure to effector functions. Front Immunol. 2014;5(OCT):1–17.

68. McManus DP, Dunne DW, Sacko M, Utzinger J, Vennervald BJ, Zhou X-N. Schistosomiasis. Nat Rev Dis Prim [Internet]. 2018;4(1):13. Available from: https://doi.org/10.1038/s41572-018-0013-8

69. Villasis E, Lopez-Perez M, Torres K, Gamboa D, Neyra V, Bendezu J, et al. Anti-Plasmodium falciparum invasion ligand antibodies in a low malaria transmission region, Loreto, Peru. Malar J [Internet]. 2012;11(1):361. Available from: https://doi.org/10.1186/1475-2875-11-361

