## Supplementary material for "Evaluation of antibody serology to determine current helminth and *Plasmodium falciparum* infections in a co-endemic area in Southern Mozambique": Table S1

**Table S1. Areas under the curve from receiver operating characteristic curve analysis for helminth-specific IgG levels and total IgE levels as markers of current infection.** Ag: Antigen; AUC: Area Under the Curve; CI: Confidence Interval; M: Microscopy; P: qPCR; MP: Microscopy and qPCR combined. Accuracy is null if AUC < 0.5, low if 0.5 ≤ AUC > 0.7, moderate if 0.7 ≤ AUC > 0.9 and high if AUC ≥ 0.9. For antigen-specific IgG, the sample size of the controls included always the 50 Spanish donors.

| **Parasite** | **Antigen** | **Method** | **Age** | **Cases** | **Controls** | **AUC** | **95% CI** | **Accuracy** |
| --- | --- | --- | --- | --- | --- | --- | --- | --- |
| *Ascaris spp.* | As16 | M | Child | 18 | 395 | 0.413 | 0.277-0.550 | Null |
|  |  |  | Adult | 15 | 387 | 0.540 | 0.371-0.708 | Low |
|  |  |  | All | 33 | 732 | 0.455 | 0.344-0.567 | Null |
|  |  | P | Child | 24 | 380 | 0.486 | 0.385-0.588 | Null |
|  |  |  | Adult | 23 | 363 | 0.587 | 0.464-0.710 | Low |
|  |  |  | All | 47 | 693 | 0.514 | 0.428-0.600 | Low |
|  |  | MP | Child | 28 | 385 | 0.450 | 0.352-0.548 | Null |
|  |  |  | Adult | 25 | 377 | 0.560 | 0.444-0.677 | Low |
|  |  |  | All | 53 | 712 | 0.483 | 0.402-0.565 | Null |
|  | As37 | M | Child | 18 | 395 | 0.322 | 0.194-0.450 | Null |
|  |  |  | Adult | 15 | 387 | 0.449 | 0.290-0.607 | Null |
|  |  |  | All | 33 | 732 | 0.372 | 0.268-0.476 | Null |
|  |  | P | Child | 24 | 380 | 0.412 | 0.308-0.515 | Null |
|  |  |  | Adult | 23 | 363 | 0.523 | 0.388-0.657 | Low |
|  |  |  | All | 47 | 693 | 0.444 | 0.355-0.532 | Null |
|  |  | MP | Child | 28 | 385 | 0.373 | 0.278-0.469 | Null |
|  |  |  | Adult | 25 | 377 | 0.530 | 0.405-0.654 | Low |
|  |  |  | All | 53 | 712 | 0.426 | 0.342-0.511 | Null |
|  | Total IgE | M | Child | 18 | 344 | 0.672 | 0.582-0.761 | Low |
|  |  |  | Adult | 15 | 336 | 0.502 | 0.333-0.672 | Low |
|  |  |  | All | 33 | 680 | 0.572 | 0.484-0.660 | Low |
|  |  | P | Child | 24 | 329 | 0.627 | 0.528-0.727 | Low |
|  |  |  | Adult | 23 | 312 | 0.505 | 0.367-0.644 | Low |
|  |  |  | All | 47 | 641 | 0.556 | 0.474-0.637 | Low |
|  |  | MP | Child | 28 | 334 | 0.626 | 0.537-0.715 | Low |
|  |  |  | Adult | 25 | 326 | 0.514 | 0.387-0.642 | Low |
|  |  |  | All | 53 | 660 | 0.558 | 0.485-0.632 | Low |
| Hookworm | AyCp2 | M | Child | 46 | 367 | 0.470 | 0.379-0.561 | Null |
|  |  |  | Adult | 72 | 330 | 0.506 | 0.432-0.580 | Low |
|  |  |  | All | 118 | 647 | 0.557 | 0.500-0.614 | Low |
|  |  | P | Child | 68 | 336 | 0.481 | 0.410-0.553 | Null |
|  |  |  | Adult | 116 | 270 | 0.490 | 0.427-0.553 | Null |
|  |  |  | All | 184 | 556 | 0.565 | 0.517-0.612 | Low |
|  |  | MP | Child | 78 | 335 | 0.468 | 0.399-0.537 | Null |
|  |  |  | Adult | 123 | 279 | 0.493 | 0.432-0.554 | Null |
|  |  |  | All | 201 | 564 | 0.554 | 0.508-0.600 | Low |
|  | NaAPR1 | M | Child | 46 | 367 | 0.519 | 0.426-0.612 | Low |
|  |  |  | Adult | 72 | 330 | 0.611 | 0.546-0.676 | Low |
|  |  |  | All | 118 | 647 | 0.592 | 0.536-0.647 | Low |
|  |  | P | Child | 68 | 336 | 0.504 | 0.427-0.581 | Low |
|  |  |  | Adult | 116 | 270 | 0.585 | 0.526-0.644 | Low |
|  |  |  | All | 184 | 556 | 0.578 | 0.531-0.625 | Low |
|  |  | MP | Child | 78 | 335 | 0.495 | 0.423-0.568 | Null |
|  |  |  | Adult | 123 | 279 | 0.588 | 0.530-0.645 | Low |
|  |  |  | All | 201 | 564 | 0.569 | 0.523-0.615 | Low |
|  | NaGST1 | M | Child | 46 | 367 | 0.547 | 0.457-0.637 | Low |
|  |  |  | Adult | 72 | 330 | 0.581 | 0.509-0.653 | Low |
|  |  |  | All | 118 | 647 | 0.581 | 0.525-0.637 | Low |
|  |  | P | Child | 68 | 336 | 0.491 | 0.412-0.571 | Null |
|  |  |  | Adult | 116 | 270 | 0.548 | 0.487-0.610 | Low |
|  |  |  | All | 184 | 556 | 0.555 | 0.507-0.604 | Low |
|  |  | MP | Child | 78 | 335 | 0.490 | 0.417-0.563 | Null |
|  |  |  | Adult | 123 | 279 | 0.546 | 0.486-0.606 | Low |
|  |  |  | All | 201 | 564 | 0.546 | 0.499-0.592 | Low |
|  | NaSAA2 | M | Child | 46 | 367 | 0.531 | 0.443-0.618 | Low |
|  |  |  | Adult | 72 | 330 | 0.585 | 0.514-0.656 | Low |
|  |  |  | All | 118 | 647 | 0.591 | 0.536-0.646 | Low |
|  |  | P | Child | 68 | 336 | 0.461 | 0.386-0.536 | Null |
|  |  |  | Adult | 116 | 270 | 0.531 | 0.469-0.594 | Low |
|  |  |  | All | 184 | 556 | 0.556 | 0.508-0.605 | Low |
|  |  | MP | Child | 78 | 335 | 0.475 | 0.404-0.546 | Null |
|  |  |  | Adult | 123 | 279 | 0.536 | 0.475-0.596 | Low |
|  |  |  | All | 201 | 564 | 0.554 | 0.508-0.601 | Low |
|  | Total IgE | M | Child | 46 | 316 | 0.701 | 0.615-0.787 | Moderate |
|  |  |  | Adult | 72 | 279 | 0.681 | 0.612-0.750 | Low |
|  |  |  | All | 118 | 595 | 0.700 | 0.648-0.752 | Moderate |
|  |  | P | Child | 68 | 285 | 0.645 | 0.574-0.715 | Low |
|  |  |  | Adult | 116 | 219 | 0.634 | 0.573-0.694 | Low |
|  |  |  | All | 184 | 504 | 0.660 | 0.616-0.704 | Low |
|  |  | MP | Child | 78 | 284 | 0.655 | 0.589-0.722 | Low |
|  |  |  | Adult | 123 | 228 | 0.633 | 0.574-0.692 | Low |
|  |  |  | All | 201 | 512 | 0.660 | 0.617-0.703 | Low |
| *Trichuris spp.* | Tm16 | M | Child | 32 | 381 | 0.548 | 0.447-0.649 | Low |
|  |  |  | Adult | 23 | 379 | 0.466 | 0.330-0.602 | Null |
|  |  |  | All | 55 | 710 | 0.473 | 0.392-0.553 | Null |
|  |  | P | Child | 64 | 340 | 0.495 | 0.422-0.567 | Null |
|  |  |  | Adult | 30 | 356 | 0.554 | 0.438-0.670 | Low |
|  |  |  | All | 94 | 646 | 0.446 | 0.383-0.508 | Null |
|  |  | MP | Child | 70 | 343 | 0.493 | 0.424-0.563 | Null |
|  |  |  | Adult | 33 | 369 | 0.536 | 0.425-0.647 | Low |
|  |  |  | All | 103 | 662 | 0.438 | 0.379-0.497 | Null |
|  | TmWAP | M | Child | 32 | 381 | 0.589 | 0.467-0.711 | Low |
|  |  |  | Adult | 23 | 379 | 0.559 | 0.438-0.679 | Low |
|  |  |  | All | 55 | 710 | 0.576 | 0.489-0.663 | Low |
|  |  | P | Child | 64 | 340 | 0.543 | 0.460-0.626 | Low |
|  |  |  | Adult | 30 | 356 | 0.532 | 0.430-0.634 | Low |
|  |  |  | All | 94 | 646 | 0.508 | 0.442-0.574 | Low |
|  |  | MP | Child | 70 | 343 | 0.556 | 0.478-0.635 | Low |
|  |  |  | Adult | 33 | 369 | 0.530 | 0.429-0.631 | Low |
|  |  |  | All | 103 | 662 | 0.511 | 0.447-0.574 | Low |
|  | Total IgE | M | Child | 32 | 330 | 0.511 | 0.404-0.618 | Low |
|  |  |  | Adult | 23 | 328 | 0.417 | 0.290-0.545 | Null |
|  |  |  | All | 55 | 658 | 0.457 | 0.378-0.536 | Null |
|  |  | P | Child | 64 | 289 | 0.528 | 0.450-0.606 | Low |
|  |  |  | Adult | 30 | 305 | 0.503 | 0.387-0.619 | Low |
|  |  |  | All | 94 | 594 | 0.489 | 0.427-0.551 | Null |
|  |  | MP | Child | 70 | 292 | 0.534 | 0.460-0.609 | Low |
|  |  |  | Adult | 33 | 318 | 0.483 | 0.373-0.593 | Null |
|  |  |  | All | 103 | 610 | 0.486 | 0.427-0.545 | Null |
| *S. stercoralis* | NIE | M | Child | 3 | 410 | 0.725 | 0.238-1.000 | Moderate |
|  |  |  | Adult | 5 | 397 | 0.896 | 0.836-0.957 | Moderate |
|  |  |  | All | 8 | 757 | 0.830 | 0.632-1.000 | Moderate |
|  |  | P | Child | 13 | 391 | 0.867 | 0.754-0.979 | Moderate |
|  |  |  | Adult | 58 | 328 | 0.836 | 0.785-0.888 | Moderate |
|  |  |  | All | 71 | 669 | 0.872 | 0.831-0.913 | Moderate |
|  |  | MP | Child | 15 | 398 | 0.834 | 0.704-0.964 | Moderate |
|  |  |  | Adult | 59 | 343 | 0.839 | 0.788-0.890 | Moderate |
|  |  |  | All | 74 | 691 | 0.865 | 0.821-0.910 | Moderate |
|  | Total IgE | M | Child | 3 | 359 | 0.786 | 0.525-1.000 | Moderate |
|  |  |  | Adult | 5 | 346 | 0.688 | 0.335-1.000 | Low |
|  |  |  | All | 8 | 705 | 0.722 | 0.487-0.956 | Moderate |
|  |  | P | Child | 13 | 340 | 0.584 | 0.461-0.708 | Low |
|  |  |  | Adult | 58 | 277 | 0.649 | 0.560-0.737 | Low |
|  |  |  | All | 71 | 617 | 0.664 | 0.590-0.738 | Low |
|  |  | MP | Child | 15 | 347 | 0.607 | 0.488-0.727 | Low |
|  |  |  | Adult | 59 | 292 | 0.637 | 0.548-0.725 | Low |
|  |  |  | All | 74 | 639 | 0.655 | 0.581-0.729 | Low |
| *Schistosoma spp.* | MEA | M | Child | 7 | 406 | 0.771 | 0.574-0.967 | Moderate |
|  |  |  | Adult | 25 | 377 | 0.569 | 0.470-0.669 | Low |
|  |  |  | All | 32 | 733 | 0.691 | 0.611-0.771 | Low |
|  |  | P | Child | 14 | 390 | 0.355 | 0.172-0.539 | Null |
|  |  |  | Adult | 57 | 329 | 0.562 | 0.487-0.637 | Low |
|  |  |  | All | 71 | 669 | 0.626 | 0.554-0.699 | Low |
|  |  | MP | Child | 19 | 394 | 0.443 | 0.281-0.605 | Null |
|  |  |  | Adult | 67 | 335 | 0.551 | 0.483-0.620 | Low |
|  |  |  | All | 86 | 679 | 0.625 | 0.562-0.688 | Low |
|  | Sm25 | M | Child | 7 | 406 | 0.639 | 0.443-0.835 | Low |
|  |  |  | Adult | 25 | 377 | 0.681 | 0.571-0.791 | Low |
|  |  |  | All | 32 | 733 | 0.721 | 0.634-0.807 | Moderate |
|  |  | P | Child | 14 | 390 | 0.484 | 0.291-0.678 | Null |
|  |  |  | Adult | 57 | 329 | 0.694 | 0.613-0.775 | Low |
|  |  |  | All | 71 | 669 | 0.712 | 0.641-0.783 | Moderate |
|  |  | MP | Child | 19 | 394 | 0.502 | 0.349-0.655 | Low |
|  |  |  | Adult | 67 | 335 | 0.671 | 0.596-0.746 | Low |
|  |  |  | All | 86 | 679 | 0.691 | 0.628-0.755 | Low |
|  | Total IgE | M | Child | 7 | 355 | 0.610 | 0.362-0.857 | Low |
|  |  |  | Adult | 25 | 326 | 0.695 | 0.598-0.792 | Low |
|  |  |  | All | 32 | 681 | 0.715 | 0.625-0.804 | Moderate |
|  |  | P | Child | 14 | 339 | 0.590 | 0.448-0.732 | Low |
|  |  |  | Adult | 57 | 278 | 0.575 | 0.489-0.660 | Low |
|  |  |  | All | 71 | 617 | 0.615 | 0.544-0.686 | Low |
|  |  | MP | Child | 19 | 343 | 0.573 | 0.443-0.703 | Low |
|  |  |  | Adult | 67 | 284 | 0.590 | 0.513-0.667 | Low |
|  |  |  | All | 86 | 627 | 0.621 | 0.557-0.685 | Low |
| *S. haematobium* | MEA | M | Child | 5 | 408 | 0.712 | 0.454-0.971 | Moderate |
|  |  |  | Adult | 11 | 391 | 0.482 | 0.364-0.600 | Null |
|  |  |  | All | 16 | 749 | 0.618 | 0.523-0.713 | Low |
|  | Sm25 | M | Child | 5 | 408 | 0.649 | 0.421-0.877 | Low |
|  |  |  | Adult | 11 | 391 | 0.714 | 0.577-0.851 | Moderate |
|  |  |  | All | 16 | 749 | 0.711 | 0.591-0.831 | Moderate |
|  | Total IgE | M | Child | 5 | 357 | 0.658 | 0.332-0.983 | Low |
|  |  |  | Adult | 11 | 340 | 0.707 | 0.546-0.869 | Moderate |
|  |  |  | All | 16 | 697 | 0.712 | 0.573-0.852 | Moderate |
| *S. mansoni* | MEA | M | Child | 2 | 411 | 0.909 | 0.734-1.000 | High |
|  |  |  | Adult | 16 | 386 | 0.612 | 0.484-0.740 | Low |
|  |  |  | All | 18 | 747 | 0.746 | 0.638-0.854 | Moderate |
|  | Sm25 | M | Child | 2 | 411 | 0.611 | 0.090-1.000 | Low |
|  |  |  | Adult | 16 | 386 | 0.665 | 0.519-0.812 | Low |
|  |  |  | All | 18 | 747 | 0.741 | 0.626-0.855 | Moderate |
|  | Total IgE | M | Child | 2 | 360 | 0.488 | 0.112-0.863 | Null |
|  |  |  | Adult | 16 | 335 | 0.695 | 0.584-0.805 | Low |
|  |  |  | All | 18 | 695 | 0.728 | 0.622-0.833 | Moderate |
| Helminths | Total IgE | M | Child | 83 | 279 | 0.658 | 0.589-0.728 | Low |
|  |  |  | Adult | 116 | 235 | 0.625 | 0.562-0.688 | Low |
|  |  |  | All | 199 | 514 | 0.649 | 0.603-0.694 | Low |
|  |  | P | Child | 149 | 204 | 0.611 | 0.552-0.671 | Low |
|  |  |  | Adult | 205 | 130 | 0.636 | 0.576-0.696 | Low |
|  |  |  | All | 354 | 334 | 0.638 | 0.597-0.679 | Low |
|  |  | MP | Child | 163 | 199 | 0.624 | 0.566-0.682 | Low |
|  |  |  | Adult | 220 | 131 | 0.627 | 0.568-0.686 | Low |
|  |  |  | All | 383 | 330 | 0.638 | 0.598-0.679 | Low |
