## Supplementary material for "Evaluation of antibody serology to determine current helminth and *Plasmodium falciparum* infections in a co-endemic area in Southern Mozambique": Table S2

**Table S2. Areas under the curve from receiver operating characteristic curve analysis for *Plasmodium falciparum*-specific IgG levels and total IgE levels as markers of current infection.** Ag: Antigen; AUC: Area Under the Curve; CI: Confidence Interval; M: Microscopy; P: qPCR; MP: Microscopy and/or qPCR. Accuracy is null if AUC < 0.5, low if 0.5 ≤ AUC > 0.7, moderate if 0.7 ≤ AUC > 0.9 and high if AUC ≥ 0.9.

| **Antigen** | **Method** | **Age** | **Cases** | **Controls** | **AUC** | **95% CI** | **Accuracy** |
| --- | --- | --- | --- | --- | --- | --- | --- |
| α-gal | P | Child | 32 | 381 | 0.504 | 0.385-0.622 | Low |
|  |  | Adult | 39 | 363 | 0.586 | 0.497-0.676 | Low |
|  |  | All | 71 | 694 | 0.547 | 0.473-0.620 | Low |
| CelTOS | P | Child | 32 | 381 | 0.592 | 0.488-0.696 | Low |
|  |  | Adult | 39 | 363 | 0.711 | 0.628-0.795 | Moderate |
|  |  | All | 71 | 694 | 0.653 | 0.581-0.725 | Low |
| SSP2 | P | Child | 32 | 381 | 0.650 | 0.540-0.759 | Low |
|  |  | Adult | 39 | 363 | 0.661 | 0.576-0.746 | Low |
|  |  | All | 71 | 694 | 0.667 | 0.599-0.735 | Low |
| LSA1 | P | Child | 32 | 381 | 0.698 | 0.597-0.799 | Low |
|  |  | Adult | 39 | 363 | 0.747 | 0.672-0.823 | Moderate |
|  |  | All | 71 | 694 | 0.707 | 0.641-0.772 | Moderate |
| EXP1 | P | Child | 32 | 381 | 0.918 | 0.883-0.953 | High |
|  |  | Adult | 39 | 363 | 0.772 | 0.702-0.842 | Moderate |
|  |  | All | 71 | 694 | 0.798 | 0.756-0.840 | Moderate |
| AMA1 | P | Child | 32 | 381 | 0.908 | 0.871-0.945 | High |
|  |  | Adult | 39 | 363 | 0.691 | 0.620-0.763 | Low |
|  |  | All | 71 | 694 | 0.785 | 0.743-0.826 | Moderate |
| EBA175 | P | Child | 32 | 381 | 0.788 | 0.709-0.867 | Moderate |
|  |  | Adult | 39 | 363 | 0.683 | 0.605-0.761 | Low |
|  |  | All | 71 | 694 | 0.721 | 0.665-0.777 | Moderate |
| MSP1 block 2 | P | Child | 32 | 381 | 0.659 | 0.551-0.767 | Low |
|  |  | Adult | 39 | 363 | 0.745 | 0.666-0.824 | Moderate |
|  |  | All | 71 | 694 | 0.685 | 0.614-0.756 | Low |
| MSP1_42_ | P | Child | 32 | 381 | 0.847 | 0.787-0.907 | Moderate |
|  |  | Adult | 39 | 363 | 0.782 | 0.715-0.848 | Moderate |
|  |  | All | 71 | 694 | 0.790 | 0.743-0.837 | Moderate |
| MSP2 | P | Child | 32 | 381 | 0.916 | 0.877-0.954 | High |
|  |  | Adult | 39 | 363 | 0.802 | 0.728-0.876 | Moderate |
|  |  | All | 71 | 694 | 0.826 | 0.783-0.869 | Moderate |
| MSP3 | P | Child | 32 | 381 | 0.839 | 0.757-0.921 | Moderate |
|  |  | Adult | 39 | 363 | 0.754 | 0.677-0.831 | Moderate |
|  |  | All | 71 | 694 | 0.772 | 0.717-0.827 | Moderate |
| MSP5 | P | Child | 32 | 381 | 0.830 | 0.759-0.901 | Moderate |
|  |  | Adult | 39 | 363 | 0.776 | 0.710-0.843 | Moderate |
|  |  | All | 71 | 694 | 0.784 | 0.733-0.835 | Moderate |
| P41 | P | Child | 32 | 381 | 0.725 | 0.625-0.825 | Moderate |
|  |  | Adult | 39 | 363 | 0.652 | 0.567-0.736 | Low |
|  |  | All | 71 | 694 | 0.689 | 0.627-0.751 | Low |
| RH1 | P | Child | 32 | 381 | 0.742 | 0.657-0.826 | Moderate |
|  |  | Adult | 39 | 363 | 0.747 | 0.675-0.818 | Moderate |
|  |  | All | 71 | 694 | 0.726 | 0.668-0.785 | Moderate |
| RH5 | P | Child | 32 | 381 | 0.632 | 0.521-0.743 | Low |
|  |  | Adult | 39 | 363 | 0.677 | 0.597-0.757 | Low |
|  |  | All | 71 | 694 | 0.653 | 0.584-0.722 | Low |
| PTRAMP | P | Child | 32 | 381 | 0.745 | 0.656-0.834 | Moderate |
|  |  | Adult | 39 | 363 | 0.754 | 0.683-0.826 | Moderate |
|  |  | All | 71 | 694 | 0.735 | 0.676-0.794 | Moderate |
| Total IgE | P | Child | 31 | 331 | 0.576 | 0.459-0.692 | Low |
|  |  | Adult | 39 | 312 | 0.595 | 0.502-0.687 | Low |
|  |  | All | 70 | 643 | 0.596 | 0.525-0.667 | Low |
