## Supplementary material for "Evaluation of antibody serology to determine current helminth and *Plasmodium falciparum* infections in a co-endemic area in Southern Mozambique": Table S3

**Table S3. Areas under the curve from receiver operating characteristic curve analysis for antigen-specific IgG levels and total IgE levels as markers of current infection, using samples without co-infections (cases) and infections (controls).** AUC: Area Under the Curve; M: Microscopy; P: qPCR; MP: Microscopy and/or qPCR. Accuracy is null if AUC < 0.5, low if 0.5 ≤ AUC > 0.7, moderate if 0.7 ≤ AUC > 0.9 and high if AUC ≥ 0.9. *MSP1 block 2.

| **Parasite** | **Antigen** | **Method** | **Cases** | **Controls** | **AUC** | **95% CI** | **Accuracy** |
| --- | --- | --- | --- | --- | --- | --- | --- |
| *Ascaris* spp. | As16 | M | 15 | 521 | 0.399 | 0.238-0.560 | Null |
|  |  | P | 16 | 357 | 0.516 | 0.387-0.645 | Low |
|  |  | MP | 18 | 352 | 0.502 | 0.371-0.633 | Low |
|  | As37 | M | 15 | 521 | 0.380 | 0.225-0.536 | Null |
|  |  | P | 16 | 357 | 0.477 | 0.339-0.614 | Null |
|  |  | MP | 18 | 352 | 0.470 | 0.334-0.607 | Null |
|  | Total IgE | M | 15 | 470 | 0.602 | 0.498-0.706 | Low |
|  |  | P | 16 | 306 | 0.570 | 0.454-0.687 | Low |
|  |  | MP | 18 | 301 | 0.581 | 0.474-0.687 | Low |
| Hookworm | AyCp2 | M | 74 | 521 | 0.542 | 0.474-0.611 | Low |
|  |  | P | 104 | 357 | 0.527 | 0.465-0.590 | Low |
|  |  | MP | 103 | 352 | 0.524 | 0.461-0.587 | Low |
|  | NaAPR1 | M | 74 | 521 | 0.590 | 0.521-0.659 | Low |
|  |  | P | 104 | 357 | 0.579 | 0.516-0.642 | Low |
|  |  | MP | 103 | 352 | 0.577 | 0.513-0.641 | Low |
|  | NaGST1 | M | 74 | 521 | 0.572 | 0.502-0.642 | Low |
|  |  | P | 104 | 357 | 0.566 | 0.501-0.631 | Low |
|  |  | MP | 103 | 352 | 0.569 | 0.504-0.634 | Low |
|  | NaSAA2 | M | 74 | 521 | 0.577 | 0.509-0.645 | Low |
|  |  | P | 104 | 357 | 0.541 | 0.477-0.605 | Low |
|  |  | MP | 103 | 352 | 0.544 | 0.480-0.609 | Low |
|  | Total IgE | M | 74 | 470 | 0.707 | 0.642-0.773 | Moderate |
|  |  | P | 104 | 306 | 0.662 | 0.602-0.721 | Low |
|  |  | MP | 103 | 301 | 0.653 | 0.592-0.713 | Low |
| *Trichuris* spp. | Tm16 | M | 30 | 521 | 0.435 | 0.328-0.542 | Null |
|  |  | P | 56 | 357 | 0.418 | 0.339-0.497 | Null |
|  |  | MP | 57 | 352 | 0.408 | 0.329-0.488 | Null |
|  | TmWAP | M | 30 | 521 | 0.614 | 0.497-0.731 | Low |
|  |  | P | 56 | 357 | 0.465 | 0.380-0.549 | Null |
|  |  | MP | 57 | 352 | 0.470 | 0.386-0.553 | Null |
|  | Total IgE | M | 30 | 470 | 0.466 | 0.359-0.573 | Null |
|  |  | P | 56 | 306 | 0.494 | 0.414-0.573 | Null |
|  |  | MP | 57 | 301 | 0.495 | 0.416-0.575 | Null |
| *S. stercoralis* | NIE | M | 3 | 521 | 0.962 | 0.925-0.998 | High |
|  |  | P | 30 | 357 | 0.910 | 0.859-0.96 | High |
|  |  | MP | 29 | 352 | 0.913 | 0.861-0.965 | High |
|  | Total IgE | M | 3 | 470 | 0.604 | 0.015-1 | Low |
|  |  | P | 30 | 306 | 0.615 | 0.485-0.745 | Low |
|  |  | MP | 29 | 301 | 0.600 | 0.465-0.735 | Low |
| *Schistosoma* spp. | MEA | M | 18 | 521 | 0.683 | 0.554-0.812 | Low |
|  |  | P | 30 | 357 | 0.600 | 0.471-0.730 | Low |
|  |  | MP | 38 | 352 | 0.624 | 0.519-0.730 | Low |
|  | Sm25 | M | 18 | 521 | 0.715 | 0.600-0.829 | Moderate |
|  |  | P | 30 | 357 | 0.669 | 0.553-0.785 | Low |
|  |  | MP | 38 | 352 | 0.690 | 0.595-0.786 | Low |
|  | Total IgE | M | 18 | 470 | 0.685 | 0.559-0.810 | Low |
|  |  | P | 30 | 306 | 0.643 | 0.533-0.752 | Low |
|  |  | MP | 38 | 301 | 0.640 | 0.542-0.738 | Low |
| *S. haematobium* | MEA | M | 10 | 521 | 0.612 | 0.466-0.758 | Low |
|  | Sm25 | M | 10 | 521 | 0.675 | 0.518-0.832 | Low |
|  | Total IgE | M | 10 | 470 | 0.639 | 0.454-0.824 | Low |
| *S. mansoni* | MEA | M | 8 | 521 | 0.772 | 0.554-0.991 | Moderate |
|  | Sm25 | M | 8 | 521 | 0.765 | 0.597-0.933 | Moderate |
|  | Total IgE | M | 8 | 470 | 0.741 | 0.580-0.903 | Moderate |
| Helminths | Total IgE | M | 173 | 470 | 0.643 | 0.594-0.692 | Low |
|  |  | P | 316 | 306 | 0.631 | 0.588-0.675 | Low |
|  |  | MP | 342 | 301 | 0.630 | 0.588-0.673 | Low |
| *P. falciparum* | α-gal | P | 30 | 352 | 0.486 | 0.371-0.601 | Null |
|  | CelTOS | P | 30 | 352 | 0.662 | 0.545-0.778 | Low |
|  | SSP2 | P | 30 | 352 | 0.628 | 0.520-0.735 | Low |
|  | LSA1 | P | 30 | 352 | 0.722 | 0.623-0.821 | Moderate |
|  | EXP1 | P | 30 | 352 | 0.828 | 0.773-0.883 | Moderate |
|  | AMA1 | P | 30 | 352 | 0.837 | 0.789-0.884 | Moderate |
|  | EBA175 | P | 30 | 352 | 0.726 | 0.639-0.812 | Moderate |
|  | MSP1* | P | 30 | 352 | 0.706 | 0.599-0.813 | Moderate |
|  | MSP1_42_ | P | 30 | 352 | 0.825 | 0.764-0.885 | Moderate |
|  | MSP2 | P | 30 | 352 | 0.855 | 0.797-0.914 | Moderate |
|  | MSP3 | P | 30 | 352 | 0.807 | 0.733-0.882 | Moderate |
|  | MSP5 | P | 30 | 352 | 0.784 | 0.709-0.859 | Moderate |
|  | P41 | P | 30 | 352 | 0.671 | 0.581-0.761 | Low |
|  | RH1 | P | 30 | 352 | 0.701 | 0.613-0.790 | Moderate |
|  | RH5 | P | 30 | 352 | 0.673 | 0.571-0.774 | Low |
|  | PTRAMP | P | 30 | 352 | 0.764 | 0.685-0.842 | Moderate |
|  | Total IgE | P | 29 | 301 | 0.551 | 0.436-0.666 | Low |
