## Supplementary material for "Evaluation of antibody serology to determine current helminth and *Plasmodium falciparum* infections in a co-endemic area in Southern Mozambique": Table S4

**Table S4. Specificity and sensitivity of seropositivity cutoffs calculated by different methods to detect current infection.** Ag: Antigen; EM: Expectation-Maximization; NC: Negative Controls; SP: Specificity; SE: Sensitivity, C: Cutoff; M: Microscopy, P: qPCR; MP: Microscopy and/or qPCR. *MSP1 block 2.

|  |  |  |  |  | **EM** | | | **NC** | | |  | **Priority 95 SP** | | | **Priority 95 SE** | | | **Youden's Index** | | |
| --- | --- | --- | --- | --- | --- | --- | --- | --- | --- | --- | --- | --- | --- | --- | --- | --- | --- | --- | --- | --- |
| **Parasite** | **Ag** | **Method** | **Cases** | **Controls** | **SP** | **SE** | **C** | **SP** | **SE** | **C** | **AUC** | **SP** | **SE** | **C** | **SP** | **SE** | **C** | **SP** | **SE** | **C** |
| *Ascaris* spp. | As16 | M | 33 | 732 | 81.69 | 21.21 | 4.00 | 95.90 | 6.06 | 4.20 | 0.455 | 95.08 | 12.12 | 4.17 | 0.82 | 96.97 | 3.75 | 92.90 | 15.15 | 4.13 |
|  |  | P | 47 | 693 | 82.54 | 21.28 | 4.00 | 96.25 | 6.38 | 4.20 | 0.514 | 95.09 | 12.77 | 4.16 | 11.40 | 95.74 | 3.77 | 15.87 | 93.62 | 3.78 |
|  |  | MP | 53 | 712 | 81.60 | 18.87 | 4.00 | 95.93 | 5.66 | 4.20 | 0.483 | 95.08 | 11.32 | 4.17 | 7.44 | 96.23 | 3.76 | 95.37 | 11.32 | 4.18 |
|  | As37 | M | 33 | 732 | 96.99 | 3.03 | 4.56 | 89.21 | 6.06 | 4.45 | 0.372 | 95.08 | 3.03 | 4.52 | 2.05 | 96.97 | 4.08 | 99.59 | 3.03 | 4.79 |
|  |  | P | 47 | 693 | 97.26 | 4.26 | 4.56 | 89.32 | 8.51 | 4.45 | 0.444 | 95.09 | 4.26 | 4.52 | 3.17 | 95.74 | 4.08 | 97.98 | 4.26 | 4.62 |
|  |  | MP | 53 | 712 | 97.05 | 3.77 | 4.56 | 89.19 | 7.55 | 4.45 | 0.426 | 95.08 | 3.77 | 4.52 | 3.09 | 96.23 | 4.08 | 97.89 | 3.77 | 4.62 |
|  | Total IgE | M | 33 | 680 | 66.47 | 36.36 | 4.32 |  |  |  | 0.572 | 95.15 | 9.09 | 4.88 | 16.18 | 96.97 | 3.31 | 33.09 | 90.91 | 3.71 |
|  |  | P | 47 | 641 | 66.46 | 36.17 | 4.32 |  |  |  | 0.556 | 95.01 | 10.64 | 4.87 | 17.63 | 95.74 | 3.33 | 21.84 | 93.62 | 3.43 |
|  |  | MP | 53 | 660 | 66.36 | 33.96 | 4.32 |  |  |  | 0.558 | 95.15 | 9.43 | 4.88 | 17.73 | 96.23 | 3.33 | 21.97 | 94.34 | 3.43 |
| Hookworm | AyCp2 | M | 118 | 647 | 32.46 | 78.81 | 4.65 | 99.85 | 0.00 | 5.07 | 0.557 | 95.05 | 6.78 | 4.91 | 5.87 | 95.76 | 4.60 | 34.47 | 77.97 | 4.66 |
|  |  | P | 184 | 556 | 33.09 | 77.17 | 4.65 | 100.00 | 0.54 | 5.07 | 0.565 | 95.14 | 5.98 | 4.91 | 10.07 | 95.11 | 4.61 | 35.97 | 76.09 | 4.66 |
|  |  | MP | 201 | 564 | 33.33 | 76.62 | 4.65 | 100.00 | 0.50 | 5.07 | 0.554 | 95.04 | 5.97 | 4.91 | 7.09 | 95.52 | 4.60 | 36.17 | 75.12 | 4.66 |
|  | NaAPR1 | M | 118 | 647 | 86.24 | 23.73 | 4.35 | 98.45 | 0.00 | 4.57 | 0.592 | 95.05 | 6.78 | 4.44 | 4.64 | 95.76 | 4.23 | 54.25 | 61.86 | 4.28 |
|  |  | P | 184 | 556 | 87.05 | 20.11 | 4.35 | 98.38 | 0.00 | 4.57 | 0.578 | 95.14 | 3.80 | 4.45 | 6.83 | 95.11 | 4.23 | 60.25 | 54.35 | 4.28 |
|  |  | MP | 201 | 564 | 86.52 | 20.40 | 4.35 | 98.23 | 0.00 | 4.57 | 0.569 | 95.04 | 3.98 | 4.45 | 6.74 | 95.02 | 4.23 | 67.20 | 45.77 | 4.29 |
|  | NaGST1 | M | 118 | 647 | 97.53 | 2.54 | 4.15 | 92.89 | 11.02 | 4.04 | 0.581 | 95.05 | 8.47 | 4.06 | 7.73 | 95.76 | 3.70 | 53.94 | 61.02 | 3.85 |
|  |  | P | 184 | 556 | 97.48 | 2.72 | 4.15 | 93.53 | 9.78 | 4.04 | 0.555 | 95.14 | 6.52 | 4.06 | 3.96 | 96.20 | 3.68 | 75.36 | 35.33 | 3.92 |
|  |  | MP | 201 | 564 | 97.52 | 2.49 | 4.15 | 92.91 | 9.45 | 4.04 | 0.546 | 95.04 | 5.97 | 4.07 | 7.09 | 95.02 | 3.69 | 48.94 | 60.70 | 3.83 |
|  | NaSAA2 | M | 118 | 647 | 84.23 | 18.64 | 4.17 | 95.98 | 8.47 | 4.29 | 0.591 | 95.05 | 11.02 | 4.26 | 6.80 | 95.76 | 3.99 | 39.72 | 77.12 | 4.05 |
|  |  | P | 184 | 556 | 85.07 | 17.93 | 4.17 | 96.04 | 5.98 | 4.29 | 0.556 | 95.14 | 8.70 | 4.26 | 5.40 | 95.11 | 3.99 | 59.35 | 51.63 | 4.08 |
|  |  | MP | 201 | 564 | 84.22 | 17.41 | 4.17 | 95.92 | 6.47 | 4.29 | 0.554 | 95.04 | 7.46 | 4.26 | 5.50 | 95.02 | 3.99 | 58.87 | 51.74 | 4.08 |
|  | Total IgE | M | 118 | 595 | 71.76 | 61.02 | 4.32 |  |  |  | 0.700 | 95.13 | 16.10 | 4.84 | 17.82 | 95.76 | 3.31 | 72.44 | 61.02 | 4.33 |
|  |  | P | 184 | 504 | 72.42 | 50.54 | 4.32 |  |  |  | 0.660 | 95.04 | 10.87 | 4.84 | 20.24 | 95.11 | 3.32 | 53.37 | 74.46 | 3.95 |
|  |  | MP | 201 | 512 | 72.85 | 50.25 | 4.32 |  |  |  | 0.660 | 95.12 | 9.45 | 4.85 | 15.82 | 95.02 | 3.21 | 53.52 | 73.63 | 3.95 |
| *Trichuris* spp. | Tm16 | M | 55 | 710 | 84.79 | 16.36 | 3.93 | 96.48 | 3.64 | 4.11 | 0.473 | 95.07 | 9.09 | 4.06 | 4.23 | 96.36 | 3.67 | 94.37 | 10.91 | 4.04 |
|  |  | P | 94 | 646 | 84.83 | 13.83 | 3.93 | 96.59 | 4.26 | 4.11 | 0.446 | 95.05 | 7.45 | 4.06 | 6.35 | 95.74 | 3.67 | 94.43 | 8.51 | 4.04 |
|  |  | MP | 103 | 662 | 84.44 | 13.59 | 3.93 | 96.53 | 3.88 | 4.11 | 0.438 | 95.02 | 6.80 | 4.06 | 6.19 | 95.15 | 3.67 | 95.32 | 6.80 | 4.06 |
|  | TmWAP | M | 55 | 710 | 50.00 | 61.82 | 4.64 | 100.00 | 0.00 | 5.04 | 0.576 | 95.07 | 14.55 | 4.89 | 3.80 | 96.36 | 4.53 | 65.49 | 50.91 | 4.70 |
|  |  | P | 94 | 646 | 50.93 | 51.06 | 4.64 | 100.00 | 0.00 | 5.04 | 0.508 | 95.05 | 10.64 | 4.89 | 3.72 | 95.74 | 4.53 | 95.98 | 10.64 | 4.90 |
|  |  | MP | 103 | 662 | 51.06 | 50.49 | 4.64 | 100.00 | 0.00 | 5.04 | 0.511 | 95.02 | 10.68 | 4.89 | 5.74 | 95.15 | 4.54 | 90.63 | 16.50 | 4.85 |
|  | Total IgE | M | 55 | 658 | 65.81 | 27.27 | 4.32 |  |  |  | 0.457 | 95.14 | 3.64 | 4.89 | 1.67 | 96.36 | 2.73 | 7.90 | 94.55 | 3.07 |
|  |  | P | 94 | 594 | 65.32 | 27.66 | 4.32 |  |  |  | 0.489 | 95.12 | 4.26 | 4.89 | 5.72 | 95.74 | 3.03 | 30.13 | 73.40 | 3.66 |
|  |  | MP | 103 | 610 | 65.25 | 27.18 | 4.32 |  |  |  | 0.486 | 95.08 | 3.88 | 4.89 | 5.90 | 95.15 | 3.03 | 30.16 | 73.79 | 3.66 |
| *S. stercoralis* | NIE | M | 8 | 757 | 75.17 | 87.50 | 4.81 | 47.82 | 87.50 | 4.73 | 0.830 | 95.11 | 25.00 | 5.10 | 13.47 | 100.00 | 4.69 | 86.66 | 87.50 | 4.97 |
|  |  | P | 71 | 669 | 80.57 | 80.28 | 4.81 | 52.17 | 92.96 | 4.73 | 0.872 | 95.07 | 43.66 | 5.05 | 45.14 | 95.77 | 4.72 | 78.92 | 84.51 | 4.80 |
|  |  | MP | 74 | 691 | 80.32 | 79.73 | 4.81 | 51.66 | 91.89 | 4.73 | 0.865 | 95.08 | 47.30 | 5.04 | 39.22 | 95.95 | 4.71 | 78.73 | 83.78 | 4.80 |
|  | Total IgE | M | 8 | 705 | 66.81 | 75.00 | 4.32 |  |  |  | 0.722 | 95.04 | 25.00 | 4.88 | 1.28 | 100.00 | 2.70 | 73.76 | 75.00 | 4.43 |
|  |  | P | 71 | 617 | 69.04 | 57.75 | 4.32 |  |  |  | 0.664 | 95.14 | 19.72 | 4.84 | 7.13 | 95.77 | 3.06 | 73.91 | 54.93 | 4.40 |
|  |  | MP | 74 | 639 | 69.01 | 56.76 | 4.32 |  |  |  | 0.655 | 95.15 | 18.92 | 4.85 | 3.29 | 95.95 | 2.89 | 89.20 | 39.19 | 4.70 |
| *Schistosoma* spp. | MEA | M | 32 | 733 | 81.72 | 28.13 | 4.27 | 66.85 | 59.38 | 4.23 | 0.691 | 95.09 | 9.38 | 4.38 | 12.82 | 96.88 | 4.13 | 54.98 | 84.38 | 4.20 |
|  |  | P | 71 | 669 | 82.81 | 30.99 | 4.27 | 68.46 | 56.34 | 4.23 | 0.626 | 95.07 | 8.45 | 4.37 | 1.35 | 97.18 | 4.10 | 67.12 | 59.15 | 4.23 |
|  |  | MP | 86 | 679 | 82.47 | 27.91 | 4.27 | 68.04 | 52.33 | 4.23 | 0.625 | 95.14 | 8.14 | 4.37 | 1.33 | 97.67 | 4.10 | 56.26 | 69.77 | 4.20 |
|  | Sm25 | M | 32 | 733 | 61.80 | 71.88 | 4.64 | 96.04 | 15.63 | 4.83 | 0.721 | 95.09 | 18.75 | 4.80 | 21.96 | 96.88 | 4.58 | 63.71 | 71.88 | 4.64 |
|  |  | P | 71 | 669 | 64.57 | 73.24 | 4.64 | 97.61 | 22.54 | 4.83 | 0.712 | 95.07 | 30.99 | 4.77 | 8.37 | 95.77 | 4.57 | 64.72 | 73.24 | 4.64 |
|  |  | MP | 86 | 679 | 64.21 | 69.77 | 4.64 | 97.35 | 18.60 | 4.83 | 0.691 | 95.14 | 25.58 | 4.78 | 9.57 | 95.35 | 4.57 | 64.36 | 69.77 | 4.64 |
|  | Total IgE | M | 32 | 681 | 67.69 | 62.50 | 4.32 |  |  |  | 0.715 | 95.01 | 15.63 | 4.87 | 25.70 | 96.88 | 3.51 | 75.33 | 62.50 | 4.44 |
|  |  | P | 71 | 617 | 67.42 | 43.66 | 4.32 |  |  |  | 0.615 | 95.14 | 12.68 | 4.87 | 12.16 | 95.77 | 3.19 | 57.54 | 61.97 | 4.12 |
|  |  | MP | 86 | 627 | 67.78 | 44.19 | 4.32 |  |  |  | 0.621 | 95.06 | 10.47 | 4.87 | 12.44 | 95.35 | 3.19 | 77.67 | 41.86 | 4.47 |
| *S. haematobium* | MEA | M | 16 | 749 | 81.04 | 6.25 | 4.27 | 65.82 | 37.50 | 4.23 | 0.618 | 95.06 | 6.25 | 4.38 | 12.68 | 100.00 | 4.13 | 54.07 | 81.25 | 4.20 |
|  | Sm25 | M | 16 | 749 | 61.15 | 75.00 | 4.64 | 95.59 | 6.25 | 4.83 | 0.711 | 95.06 | 12.50 | 4.81 | 13.08 | 100.00 | 4.58 | 63.02 | 75.00 | 4.64 |
|  | Total IgE | M | 16 | 697 | 67.00 | 62.50 | 4.32 |  |  |  | 0.712 | 95.12 | 12.50 | 4.88 | 4.30 | 100.00 | 2.92 | 76.76 | 62.50 | 4.48 |
| *S. mansoni* | MEA | M | 18 | 747 | 81.93 | 44.44 | 4.27 | 66.80 | 77.78 | 4.23 | 0.746 | 95.05 | 11.11 | 4.38 | 2.54 | 100.00 | 4.11 | 69.08 | 77.78 | 4.24 |
|  | Sm25 | M | 18 | 747 | 61.18 | 72.22 | 4.64 | 95.98 | 22.22 | 4.83 | 0.741 | 95.05 | 22.22 | 4.80 | 21.69 | 100.00 | 4.58 | 84.47 | 55.56 | 4.72 |
|  | Total IgE | M | 18 | 695 | 67.19 | 66.67 | 4.32 |  |  |  | 0.728 | 95.11 | 16.67 | 4.88 | 25.32 | 100.00 | 3.51 | 74.68 | 66.67 | 4.44 |
| Helminths | Total IgE | M | 199 | 514 | 72.76 | 50.25 | 4.32 |  |  |  | 0.649 | 95.14 | 14.57 | 4.81 | 9.73 | 95.48 | 3.09 | 82.10 | 42.21 | 4.47 |
|  |  | P | 354 | 334 | 74.85 | 41.81 | 4.32 |  |  |  | 0.638 | 95.21 | 12.71 | 4.78 | 11.08 | 95.20 | 3.08 | 67.66 | 53.11 | 4.15 |
|  |  | MP | 383 | 330 | 75.15 | 41.25 | 4.32 |  |  |  | 0.638 | 95.15 | 12.79 | 4.78 | 10.30 | 95.04 | 3.07 | 46.67 | 75.20 | 3.75 |
| *P. falciparum* | α-gal | P | 71 | 694 | 92.22 | 12.68 | 4.63 | 74.35 | 33.80 | 4.48 | 0.547 | 95.10 | 4.23 | 4.72 | 7.78 | 95.77 | 4.20 | 70.75 | 43.66 | 4.46 |
|  | Celtos | P | 71 | 694 | 76.80 | 52.11 | 4.28 | 99.28 | 0.00 | 4.81 | 0.653 | 95.10 | 14.08 | 4.51 | 11.10 | 95.77 | 4.10 | 81.12 | 52.11 | 4.30 |
|  | SSP2 | P | 71 | 694 | 57.49 | 67.61 | 4.33 | 98.70 | 1.41 | 4.76 | 0.667 | 95.10 | 5.63 | 4.61 | 7.64 | 95.77 | 4.26 | 68.30 | 63.38 | 4.37 |
|  | LSA1 | P | 71 | 694 | 74.78 | 56.34 | 4.22 | 92.65 | 25.35 | 4.36 | 0.707 | 95.10 | 21.13 | 4.40 | 13.54 | 95.77 | 4.15 | 79.83 | 54.93 | 4.25 |
|  | EXP1 | P | 71 | 694 | 44.52 | 98.59 | 4.99 | 49.71 | 95.77 | 5.01 | 0.798 | 95.10 | 22.54 | 5.32 | 54.32 | 95.77 | 5.02 | 54.32 | 95.77 | 5.02 |
|  | AMA1 | P | 71 | 694 | 31.70 | 100.00 | 4.97 | 41.07 | 97.18 | 5.02 | 0.785 | 95.10 | 15.49 | 5.32 | 50.43 | 95.77 | 5.09 | 53.17 | 94.37 | 5.11 |
|  | EBA175 | P | 71 | 694 | 57.78 | 74.65 | 4.74 | 80.69 | 36.62 | 4.88 | 0.721 | 95.10 | 23.94 | 5.00 | 27.23 | 95.77 | 4.70 | 55.76 | 81.69 | 4.73 |
|  | MSP1 | P | 71 | 694 | 91.07 | 28.17 | 4.07 | 48.13 | 73.24 | 3.97 | 0.685 | 95.10 | 18.31 | 4.15 | 8.21 | 95.77 | 3.95 | 76.95 | 59.15 | 4.01 |
|  | MSP1_42_ | P | 71 | 694 | 41.07 | 95.77 | 4.98 | 41.79 | 95.77 | 4.99 | 0.790 | 95.10 | 26.76 | 5.29 | 45.68 | 95.77 | 5.00 | 63.40 | 84.51 | 5.10 |
|  | MSP2 | P | 71 | 694 | 47.98 | 95.77 | 4.95 | 41.79 | 97.18 | 4.94 | 0.826 | 95.10 | 30.99 | 5.17 | 49.86 | 95.77 | 4.95 | 75.79 | 74.65 | 5.04 |
|  | MSP3 | P | 71 | 694 | 51.44 | 88.73 | 4.93 | 80.12 | 60.56 | 5.05 | 0.772 | 95.10 | 26.76 | 5.17 | 25.79 | 95.77 | 4.89 | 58.79 | 85.92 | 4.95 |
|  | MSP5 | P | 71 | 694 | 60.66 | 85.92 | 4.65 | 76.08 | 74.65 | 4.73 | 0.784 | 95.10 | 15.49 | 4.93 | 36.46 | 95.77 | 4.60 | 76.95 | 74.65 | 4.73 |
|  | P41 | P | 71 | 694 | 73.78 | 50.70 | 4.81 | 92.22 | 18.31 | 4.92 | 0.689 | 95.10 | 16.90 | 4.95 | 21.61 | 95.77 | 4.73 | 55.19 | 76.06 | 4.77 |
|  | PfRh1 | P | 71 | 694 | 65.42 | 67.61 | 4.64 | 87.90 | 36.62 | 4.74 | 0.726 | 95.10 | 5.63 | 4.82 | 25.07 | 95.77 | 4.56 | 80.84 | 57.75 | 4.69 |
|  | PfRh5 | P | 71 | 694 | 90.92 | 16.90 | 4.26 | 83.14 | 36.62 | 4.22 | 0.653 | 95.10 | 9.86 | 4.37 | 13.98 | 95.77 | 4.15 | 63.54 | 66.20 | 4.19 |
|  | PTRAMP | P | 71 | 694 | 70.03 | 64.79 | 4.67 | 79.39 | 53.52 | 4.72 | 0.735 | 95.10 | 16.90 | 4.88 | 19.31 | 95.77 | 4.59 | 75.94 | 63.38 | 4.70 |
|  | Total IgE | P | 70 | 643 | 67.65 | 45.71 | 4.32 |  |  |  | 0.596 | 95.02 | 11.43 | 4.87 | 11.20 | 95.71 | 3.15 | 71.23 | 45.71 | 4.37 |
