## Supplementary material for "Evaluation of antibody serology to determine current helminth and *Plasmodium falciparum* infections in a co-endemic area in Southern Mozambique": Figure S1

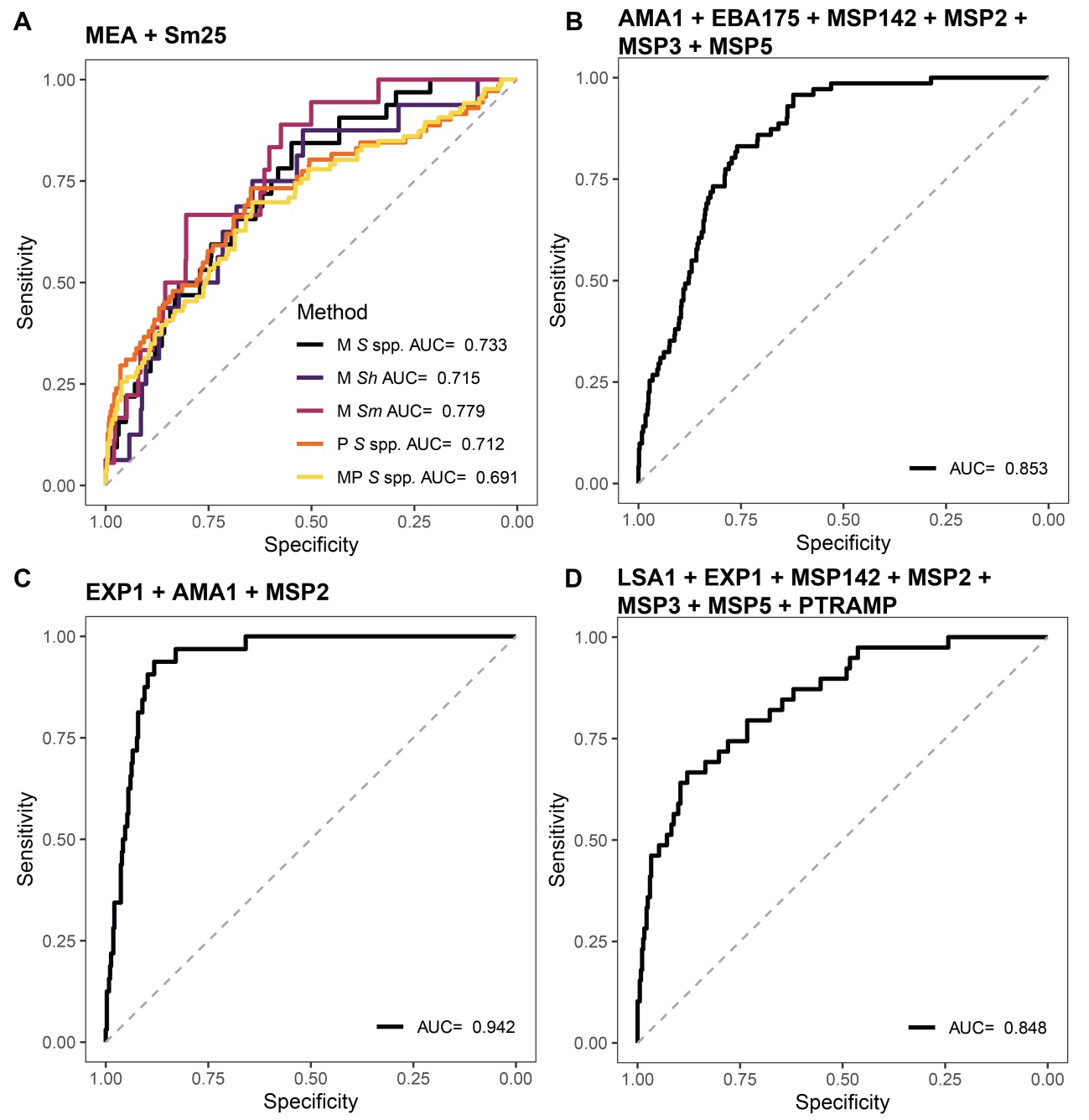

**Fig S1. Receiver operating characteristic curves and their corresponding areas under the curve from the combination of antigens from *Schistosoma* spp. and *Plasmodium falciparum*.** The predicted values fitted by regression models were used to build the Receiver operating characteristic (ROC) curves and calculate the corresponding areas under the curve (AUCs). The models were built using the combination of log_10_-transformed median fluorescence intensity (MFI) as predictor variable and the Microscopic (M), qPCR (P) or the combination of both (MP) diagnoses were used as response variables. For *Schistosoma* spp. (**A**), the combination of the MFI from the two antigens in the panel was used. For *P. falciparum* the combination of the MFI from antigens that gave the best AUC with the lowest number of predictors is represented. We included only the top performer antigens to calculate the combinations, whether it was for the whole population (**B**) or stratified by age (**C**, children; **D**, adults). To build the models, serum samples from 715 endemic individuals and 50 Spanish donors were used. *S* spp.: *Schistosoma* spp.; *Sh*: *Schistosoma haematobium*; *Sm*: *Schistosoma mansoni*.
