## Supplementary material for "Evaluation of antibody serology to determine current helminth and *Plasmodium falciparum* infections in a co-endemic area in Southern Mozambique": Figure S3

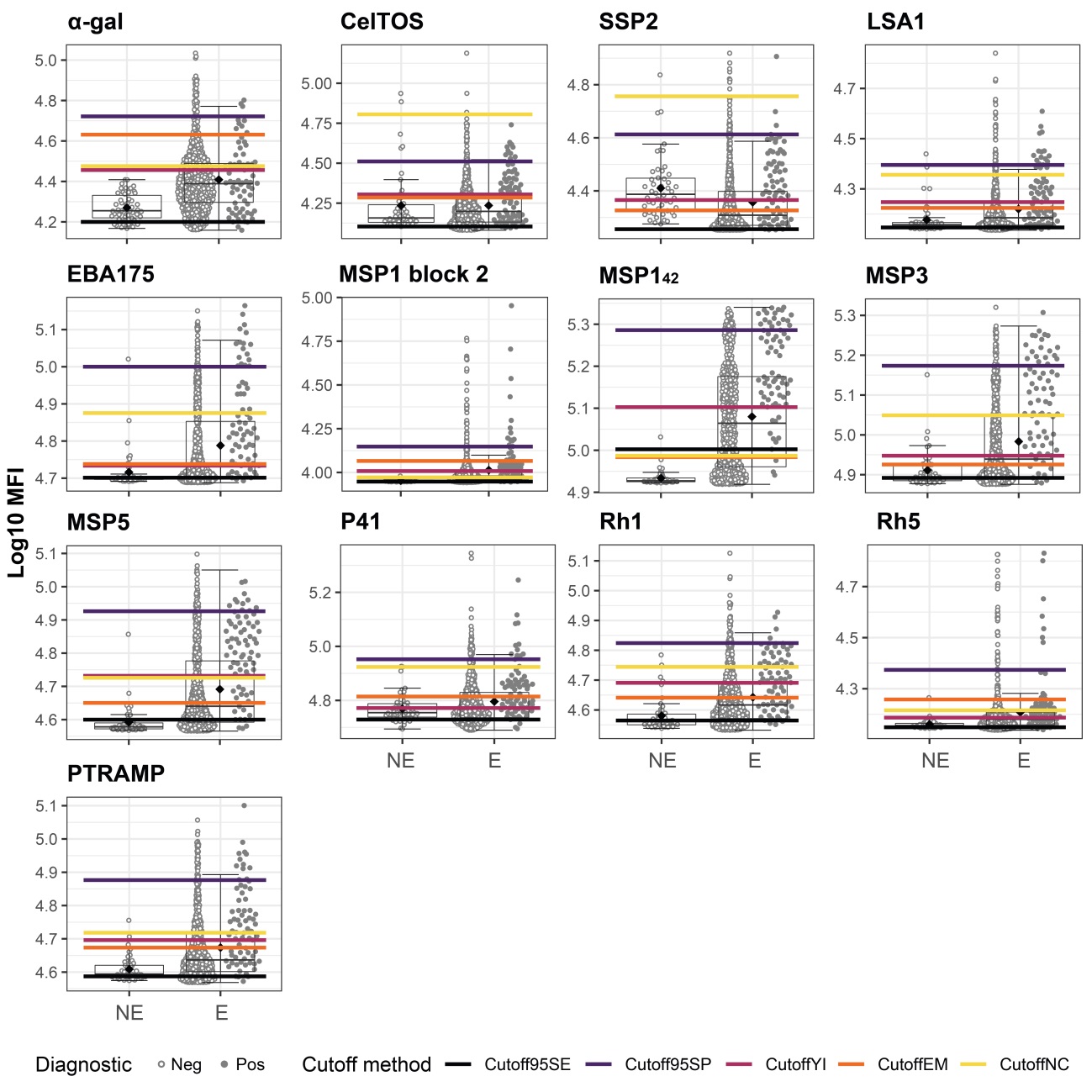

**Fig S3. *Plasmodium falciparum* antigen-specific IgG levels in non-endemic and endemic populations and seropositivity cutoffs calculated by different methods.** Antibody levels are presented as the log_10_-transformed median fluorescence intensity (MFI) of IgG against *P. falciparum* antigens. In the endemic population, filled dots highlight individuals with a *P. falciparum* infection detected by qPCR. The boxplots represent the median (bold line), the mean (black diamond), the first and third quartiles (box) and the largest and smallest values within 1.5 times the interquartile range (whiskers). Data beyond the end of the whiskers are outliers. Cutoff95SE: cutoff prioritizing a minimum sensitivity of 95%; Cutoff95SP: cutoff prioritizing a minimum specificity of 95%; CutoffYI: cutoff corresponding to the maximum Youden’s Index; CutoffEM: cutoff defined by the Expectation-Maximization algorithm; CutoffNC: cutoff calculated as median + 3*standard deviations of the log_10_-transformed MFI levels from the non-endemic population. NE: Non-Endemic (N = 50), E: Endemic (N = 715). The rest of the antigens are in Fig 8.
