## Supplementary material for "Evaluation of antibody serology to determine current helminth and *Plasmodium falciparum* infections in a co-endemic area in Southern Mozambique": Figure S4

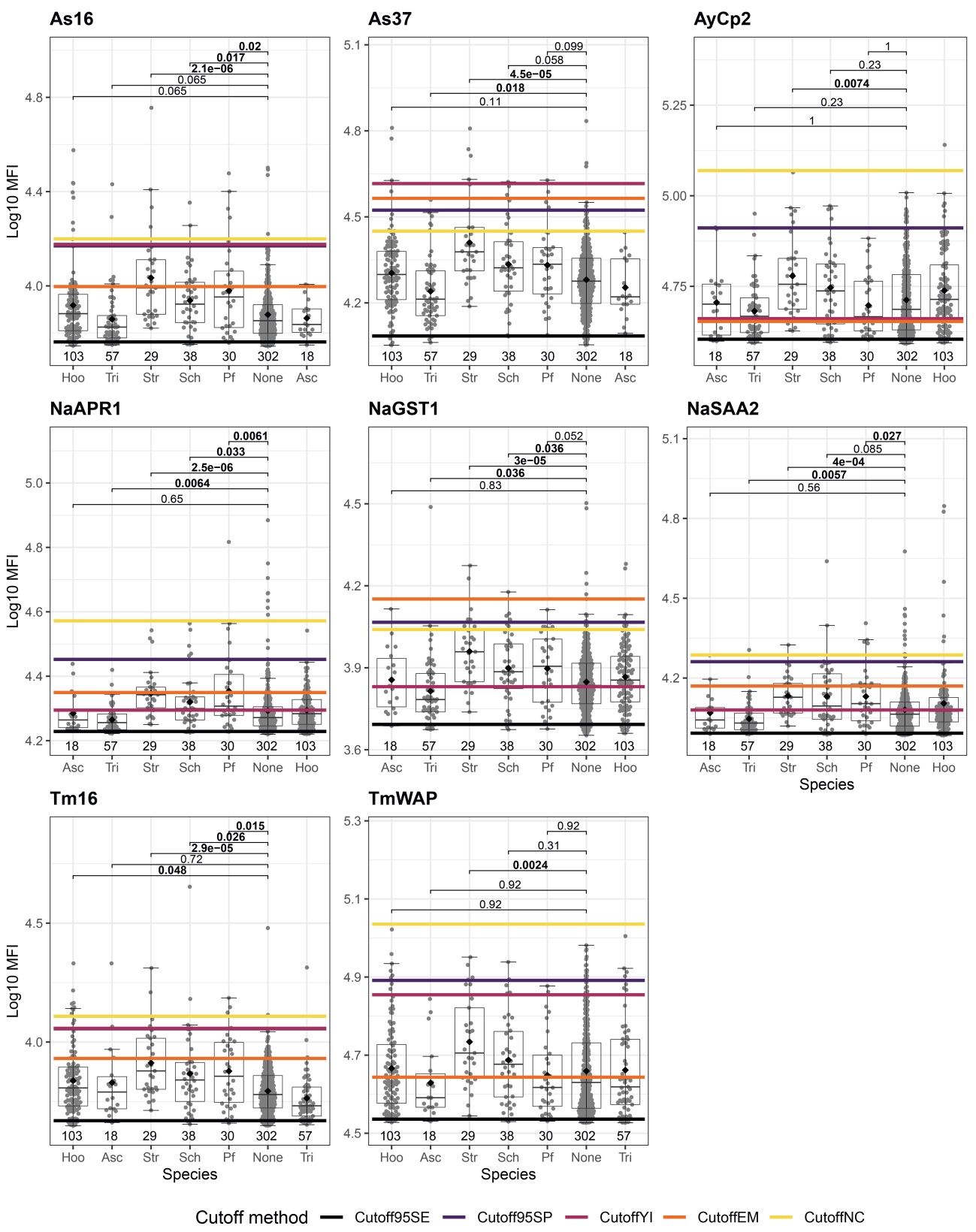

**Fig S4. Effect of single infections on IgG levels to antigens from helminths.** Only individuals with single infections or no infections at all detected by the combination of microscopy and qPCR are shown. Antibody levels are presented as the log_10_-transformed median fluorescence intensity (MFI) of IgG against helminth antigens. For each antigen, antibody levels for each species were compared to the levels of those with no infection and the corresponding species to the antigen was shown as a reference. Statistical comparison between groups was performed by Wilcoxon rank-sum test and the adjusted P values by the Holm approach are shown. Statistically significant P values are highlighted in bold. The sample size of each group is shown below each boxplot. The boxplots represent the median (bold line), the mean (black diamond), the first and third quartiles (box) and the largest and smallest values within 1.5 times the interquartile range (whiskers). Data beyond the end of the whiskers are outliers. Cutoff95SE: cutoff prioritizing a minimum sensitivity of 95%; Cutoff95SP: cutoff prioritizing a minimum specificity of 95%; CutoffYI: cutoff corresponding to the maximum Youden’s Index; CutoffEM: cutoff defined by the Expectation-Maximization algorithm; CutoffNC: cutoff calculated as median + 3*standard deviations of the log_10_-transformed MFI levels from the non-endemic population. The rest of the antigens are in Fig 9.
